# Mechanistic Heterogeneity in Type 2 Diabetes and Hypertension Comorbidity Revealed with Partitioned Polygenic Scores

**DOI:** 10.1101/2025.03.02.25323190

**Authors:** Vincent Pascat, Liudmila Zudina, Lucas Maurin, Anna Ulrich, Jared G. Maina, Ayse Demirkan, Zhanna Balkhiyarova, Igor Pupko, Yevheniya Sharhorodska, François Pattou, Bart Staels, Marika Kaakinen, Amna Khamis, Amélie Bonnefond, Patricia Munroe, Philippe Froguel, Inga Prokopenko

## Abstract

Type 2 diabetes (T2D) and hypertension are common health conditions that often occur together, suggesting shared biological mechanisms. To explore this relationship, we analysed large-scale multiomic data to uncover genetic factors underlying T2D and blood pressure (BP) comorbidity.

We curated 1,304 independent single-nucleotide variants (SNVs) associated with T2D/BP, grouping them into five clusters related to **metabolic syndrome, inverse T2D-BP risk, impaired pancreatic beta-cell function, higher adiposity, and vascular dysfunction**. Colocalisation with tissue-specific gene expression highlighted significant enrichment in pathways related to thyroid function and fetal development.

Partitioned polygenic scores (PGS) derived from these clusters improved risk prediction for T2D-hypertension comorbidity, identifying individuals with more than twice usual susceptibility.

These results reveal complex genetic basis of shared T2D and BP mechanistic heterogeneity, enhancing comorbidity risk prediction. Partitioned PGSs offer promising approach for early risk stratification, personalised prevention, and improved management of these interconnected conditions, supporting precision medicine and public health initiatives.

## Introduction

Hypertension and type 2 diabetes (T2D) pose major public health challenges, affecting approximately 1.28 billion^1^ and 537 million adults worldwide^2^, respectively, with the prevalence of T2D expected to rise to 1.3 billion by 2050. T2D and high blood pressure (BP) frequently co-occur in the same individual^3–5^. The T2D-BP comorbidity further increases the risk of major health outcomes, and individuals with both conditions often face challenges in achieving treatment objectives^6^. T2D and high BP are key part of the Metabolic Syndrome (MetS), also encompassing various cardiovascular risk factors, including central obesity, dyslipidaemia, microalbuminuria, and insulin resistance (IR)^7,8^.

Extensive genetic research, notably through recent genome-wide association studies (GWAS), has dissected the underlying genetic architecture of both T2D and BP traits independently. Latest reports associated 1,289 independent variants in DNA with T2D^9^, while 2,103 variants are implicated in BP control^10^. These findings highlight the complex genetic architecture of T2D and BP traits/hypertension, emphasising their diverse genetic drivers.

Despite significant advances in understanding the genetics of T2D and high BP as independent conditions, the shared genetic basis underlying their frequent comorbidity remains largely unexplored. This gap persists even though T2D – high BP comorbidity has been consistently observed, including within genetic datasets^11–13^. Several Mendelian randomization (MR) studies have yielded conflicting evidence about the causal relationship between T2D and high BP. For instance, Sun et al. identified T2D as driver of high BP, while Aikens et al. proposed the opposite^14,15^. Additionally, a recent analysis found that two of four types of hypertensive medications were protective against T2D risk, while the others increased its risk^16^. These findings highlight the diverse and complex pathways underpinnings the T2D-BP relationship.

In this study, we aimed to characterise the shared pathophysiological processes underlying the T2D-BP relationship by harnessing large-scale genomic datasets from recent research on both conditions. Through this approach, we sought to enhance the mechanistic understanding of these disease comorbid status and their shared genetic basis, and suggest potential avenues for targeted interventions and precision health.

We aggregated genomic data from 49 GWAS for related conditions and traits/endophenotypes, 50 tissue-specific expression quantitative trait *loci* (eQTL)^17–19^, assay for transposase-accessible chromatin using sequencing peaks from single-cell (scATAC-seq) atlas^20^, and the UK Biobank (UKB) cohort^21^. By leveraging GWAS meta-analyses summary statistics, we assessed the genetic correlation between T2D^22^ and systolic BP (SBP), diastolic BP (DBP), and pulse pressure (PP=SBP-DBP)^12^. We clustered the T2D-BP associated independent single-nucleotide variant (SNV) effects into distinct groups based on their underlying pathogenetic processes. We observed the cluster-associated changes in gene expression through colocalization analysis with eQTL and enrichment in scATAC-seq atlas. We finally evaluated the cluster-specific risks of complication using partitioned polygenic scores (PGS) in 459,247 individuals (***Figure 1***).

**Figure 1.**
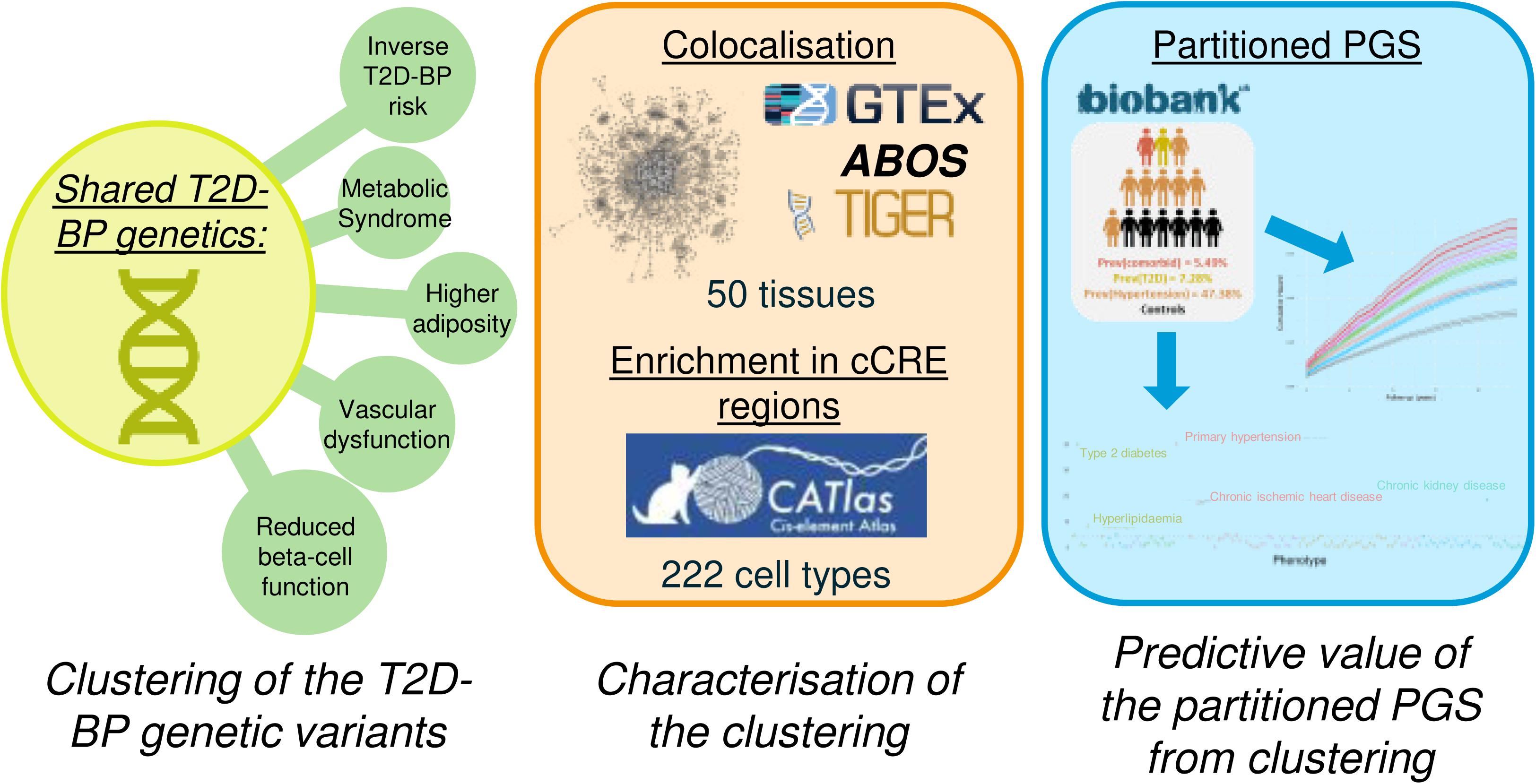
Study overview. The genetic landscape of T2D-BP is explored and subsequently clustered into five groups of different pathogenetic processes. These clusters are then analysed using colocalization methods, eQTL and the scATAC-seq atlas to identify associated changes in gene expression. The cluster are evaluated for their predictive value and the underlying causal mechanisms contributing to the T2D-BP relationship. <u>Legend:</u> T2D = type 2 diabetes; BP = blood pressure; cCRE = candidate cis-regulatory elements; PGS = polygenic scores; DBP = diastolic blood pressure; SBP = systolic blood pressure.

## Results

### Genetic overlap between T2D and BP

We explored the genetic relationships between T2D and BP by evaluating the overall genetic correlation and associated *loci* which overlap between the two conditions. We performed linkage disequilibrium (LD) score regression using *ldsc*^23^ and observed robust positive genetic correlations between T2D and SBP (r_g_[SE]=0.25[0.028], P=1.56 × 10^−19^), DBP (r_g_[SE]=0.18[0.027], P=1.38 × 10^−11^), and PP (r_g_[SE]=0.23[0.029], P=2.25 × 10^−15^).

We gathered a collection of 1,401 SNVs associated with T2D, high SBP, DBP, and/or PP (***Methods***). We constructed PGSs for T2D, SBP, DBP and PP in the UKB (***Supplementary Table 1***) using independent weights from GWASs (***Methods***). We probed whether a genetic predisposition towards one condition could predict the risk of the other using *comorbidPGS*^24^. T2D PGS was consistently associated with a modest increase in SBP, DBP, and PP (Beta_PP_[SE]≥0.37[0.017] change in PP mmHg per one-unit increase in T2D PGS, P≤1.83×10^106^). SBP and PP PGSs were significantly associated with a higher risk of T2D (OR_SBP_[95% CI]=1.07 [1.06-1.09] change in T2D odds per one-unit increase in SBP PGS, P=9.36×10^-35^; OR_PP_[95% CI]=1.07 [1.06-1.08], P=8.02×10^-31^). In contrast, DBP PGS had no impact on T2D risk (OR_DBP_[95% CI]=1.01[0.999-1.02], P=0.062, ***Supplementary Table 2***).

We revealed 24/19/26 genetic *loci* overlapping between T2D and SBP/DBP/PP respectively, determined by genomic proximity (***Supplementary Table 3***). Of the 1,401 SNVs, 9 were directly associated with both T2D and BP traits. Additionally, we identified 97 SNV pairs that overlapped (within 500 kb or LD r²>0.2) and were associated with T2D and BP traits. We observed several well-known *loci* and genes, such as those at *GRB14-COBLL1* (associated with reduced insulin level, pulse pressure, and mean arterial pressure)^25,26^, *ADCY5* (beta cell function and lipodystrophy)^27^, and *ACE* (renin-angiotensin system, hypertension)^28^. Overlapping *loci* at *JAZF1* (regulating glucose, lipid, and inflammation)^29^, *ADRB1* (beta-adrenergic receptors regulating cardiac contractility and heart rate)^30^, *TCF7L2* (controlling Langerhans islet proliferation)^31^, and *SGIP1* (signalling in energy homeostasis)^32^ contribute to the inverse relationship between T2D and BP. These results highlight the dense and complex genetic relationships between high BP predisposition and T2D risk.

### Clusters of pathogenetic processes

To dissect the complexity of shared biological pathways between T2D and BP, we curated and refined the SNV list to 1,304 independent variants (LD r²<0.2), including 500 T2D-associated and 813 BP-associated SNVs, to cluster them based on their effects on 49 related (endo)phenotypes (***Supplementary Table 4***). Our investigation encompassed a wide array of related endpoints or risk factors, including biomarkers of inflammation and hepatic function, circulating plasma lipids, cardiovascular indicators, anthropometric metrics, glycaemic traits, and sex hormones (***Supplementary Table 4***). All SNVs (originally associated with T2D, BP traits, or both) were aligned to the T2D risk allele. When information for a particular phenotype at an SNV was unavailable, we used LD proxies (***Methods***)^33^, and performed imputation of the remaining missing data by random forest algorithm implemented in the *imputeSCOPA* software tool^34^. We used an unsupervised hierarchical clustering approach, anticipating the increased heterogeneity within our SNV set, given the inherent heterogeneity in both T2D and BP signals^35^. To ensure robustness of our clustering, we ran extensive sensitivity analyses (***Methods***) with different sets of metabolic traits and other clustering methods, such as *MRClust*^36^ and Bayesian nonnegative matrix factorization (bNMF) (***Methods, Supplementary Figure 1, 2, and 3***). We identified five clusters of distinct pathogenetic mechanisms (***Figure 2, Supplementary Table 5-6***), highlighting mechanistic heterogeneity in T2D-BP comorbidity. We compared the SNV assignments of our T2D-BP clusters with recent T2D hierarchical clustering^9^ and bNMF clustering^37^ (***Methods, Supplementary Table 5, Supplementary Figure 4***). The pathophysiological processes identified across the five clusters were consistent with existing evidence and highlight novel mechanistic insights (***Methods, Supplementary Figure 5***)^9,12,37^.

**Figure 2.**
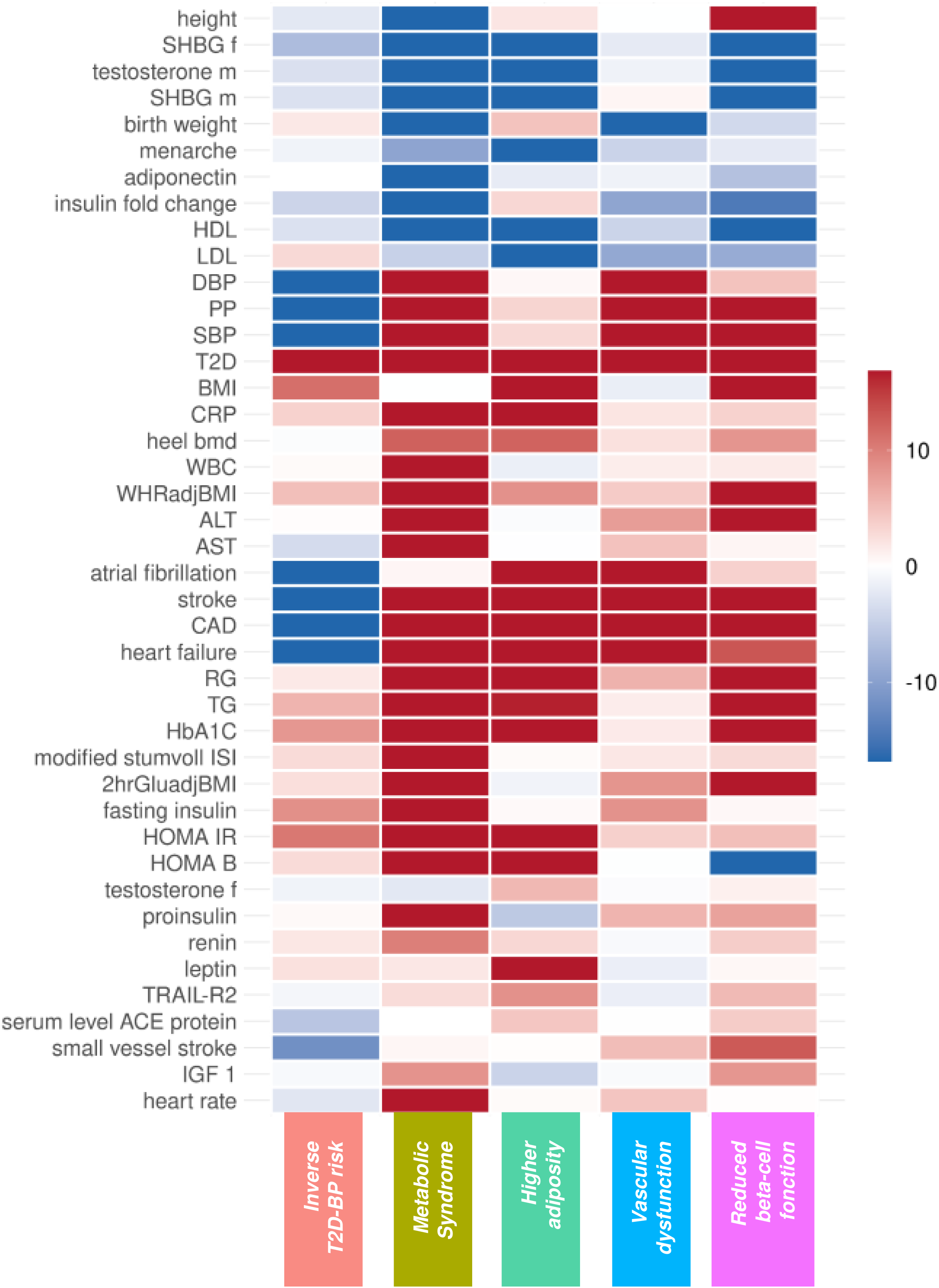
Clustering heat map of 49 endophenotypes with five pathogenetic SNV clusters associated with high BP and/or risk of T2D. Each row corresponds to a GWAS of an endophenotype, while each column corresponds to a cluster. Colour represents the direction of the z-score (aligned to the T2D risk allele) of association between the GWAS and SNVs assigned to the cluster. Colour intensity represents the associated -log(P-value). <u>Legend:</u> T2D = type 2 diabetes; DBP = diastolic blood pressure; UKB = UK Biobank; PP = pulse pressure; SBP = systolic blood pressure; HbA1C = glycated hemoglobin; RG = random glucose; WHR = waist-hip ratio; BMI = body mass index; IL = interleukin; HDL = high-density lipoprotein; PAI = plasminogen activator inhibitor; ISI = insulin sensitivity index; IGF = insulin-like growth factor; LDL = low-density cholesterol; adjBMI = adjusted for BMI; HOMA = homeostatic model assessment; IR = insulin resistance; B = beta-cell function; WBC = white blood cell count; CRP = C-reactive protein; CAD = coronary artery disease; TG = triglycerides; SHBG = sex-hormone-binding globulin; ACE = angiotensin-converting enzyme; TRAIL-R2 = TNF related apoptosis inducing ligand receptor 2; ALT = alanine aminotransferase; AST = aspartate aminotransferase; BMD = bone mass density.

### The Metabolic Syndrome cluster

included 215 variants, and displayed the most distinct pathogenetic signature. It highlights attributes consistent with the metabolic syndrome, including lower levels of sex hormones (sex-hormone binding globulin, insulin, testosterone)^38^, increased central adiposity (waist-to-hip ratio [WHR] adjusted for body-mass index [BMI])^39^, systemic higher IR evaluated by the homeostasis model assessment of insulin resistance, HOMA-IR, using both fasting plasma glucose and insulin (alongside higher HOMA-B, proinsulin level, and insulin fold change), lower high-density lipoprotein (HDL) cholesterol, higher triglycerides (TG), and altered cardiovascular functions (higher heart rate, increased cardiovascular event risk, higher renin-angiotensin-aldosterone system activity)^8^. SNVs within this cluster also strongly associated with shorter stature (lower height) and lower birth weight but do not affect overall adiposity (proxied by BMI).

Previous findings reported that shorter stature is associated with a higher risk of T2D^40^ and cardiovascular events^41^. Other studies linked greater height with insulin and insulin-like growth factor signalling pathways^42^. The impaired insulin sensitivity may be one of the underlying factors in this association^43,44^.

The T2D – high BP comorbidity is high in this cluster and was consistent with our bNMF clustering (***Supplementary Figure 3***). The origin of the SNVs is an equal mix of T2D and BP (***Supplementary Figure 1).*** When comparing SNVs with previous T2D clustering, we observed an overlap between the SNVs in our *Metabolic Syndrome* cluster and the Type 2 Diabetes Global Genetics Initiative (T2DGGI) *Metabolic syndrome* cluster, as well as the bNMF cluster *Lipodystrophy 1* (***Supplementary Figure 4***).

**In the *Inverse T2D-BP risk* cluster**, we noted an inverse relationship of associated SNVs effects on higher T2D risk related to lower SBP/DBP/PP. Predominantly originating from associations with BP traits (***Supplementary Figure 1***), the 353 SNVs within this cluster, when aligned to the T2D risk allele, are associated with a lower risk of cardiovascular events, such as atrial fibrillation (AF), coronary artery disease (CAD), stroke and heart failure. Additionally, these SNVs demonstrated associations with BMI and systemic higher IR (higher HOMA-IR).

### The Higher adiposity cluster

contained 137 SNVs – predominantly T2D signals – which showcased effects on higher BMI, reduced sex hormones (testosterone and SHBG), lower HDL- and LDL-cholesterol, higher risk of cardiovascular events (CAD, heart rate, stroke), and insulin resistance (higher HOMA-IR/HOMA-B). This cluster showed a high number of obesity-related SNVs within previous T2D clustering (***Supplementary Figure 4***). **The *Vascular Dysfunction* cluster** includes 287 SNVs mostly originating as BP signals. They are associated with cardiovascular traits (higher risk of AF, stroke, CAD, heart failure), lower birth weight and show strong effects on both T2D – high BP. Lastly, ***Reduced beta-cell function* cluster** exhibited characteristics of impaired beta-cell function including lower homeostasis model assessment of beta cell function (HOMA-B), higher glucose/glycated haemoglobin levels (random glucose [RG], HbA1c), metabolic dysregulation (TG, sex hormones), higher inflammation (C-reactive protein [CRP], IGF-1) and taller stature (height). The *Reduced beta-cell function* cluster contained 312 SNVs, predominantly T2D signals, found in the *Beta cell 1* and *Beta cell 2* clusters from the latest published T2D bNMF clustering (***Supplementary*** Figure 4b).

The five distinct mechanistic groups of genetic variants, revealed through clustering, contribute to the shared susceptibility for T2D and high BP, providing a foundation for further exploration of the biological pathways.

### Multiomic characterisation of T2D-BP clusters

To further characterise the T2D-BP comorbidity clusters, we evaluated the changes in gene expression and regulatory elements associated with the clustered SNVs. We first conducted a colocalization analysis to elucidate the impact of our SNVs on gene expression patterns. We explored the genomic landscape within a 200 kb window surrounding each clustered SNV to assess the likelihood of a shared causal variant between our clusters and gene expression changes across 50 tissues from various eQTLs datasets using a hypothesis-free approach and the *coloc* R package (**Methods**)^45^. We identified a total of 6,321 colocalizations across the 50 tissues, involving 1,558 genes and 448 clustered variants (***Figure 3a*** top***, Supplementary Table 7, Supplementary Figure 6***).

**Figure 3.**
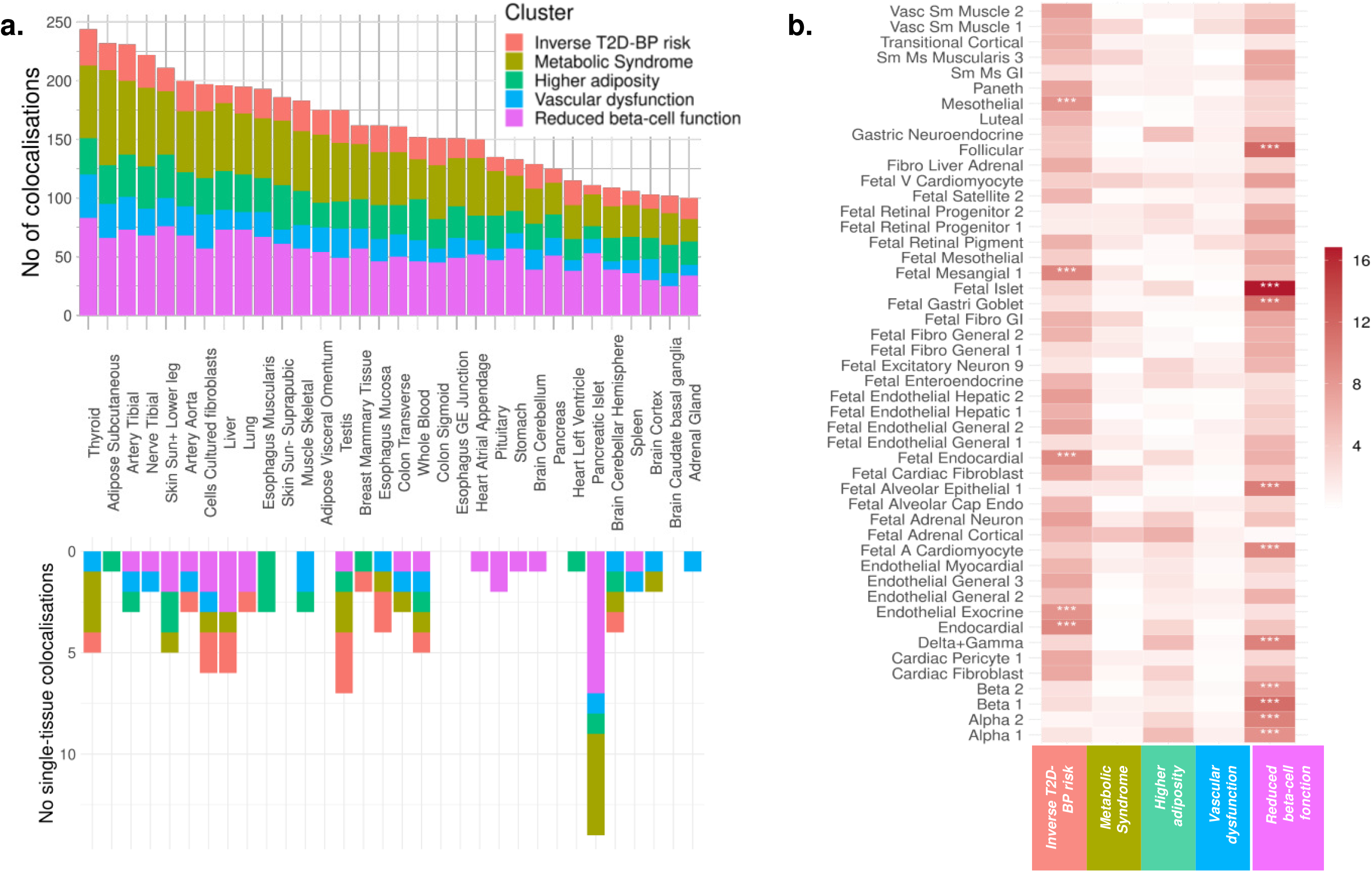
Characterisation of pathogenetic clusters: genetic expression, and regulatory mechanisms. **a**, Histogram depicting the distribution of colocalised loci across clusters in 50 human adult tissues. Each bar represents the number of colocalised signal per tissue, with colours indicating contribution from the five clusters. The upper histogram shows the overall number of colocalizations while the lower portion highlights tissue-specific colocalizations. **b**, Heat map of enrichment of open chromatin region across the five clusters and 222 cell types from 30 human adult tissues and 15 human fetal tissues. Each column corresponds to a cluster, while each row represents a cell type. Colour intensity indicates the -log(P-value) of the log fold enrichment, with asterisks denoting significant signals following Bonferroni correction. <u>Legend:</u> No = number; T2D = type 2 diabetes; BP = blood pressure; Sun+ = sun-exposed; Vasc = vascular; Sm = smooth; Ms = muscle; GI = gastrointestinal; V = ventricular; Fibro = fibroblast; A = atrial.

Our analysis revealed distinct gene expression signatures for each cluster, corroborating the diversity of the biological pathways involved. The *Inverse T2D-BP risk* cluster displays colocalization in the brain, particularly brain cerebellum (*MGRN1*, *HELLS*, *SLC39A13*) and adrenal glands (*SLC7A1*, *RHOC*, *NUDT2*). The *Metabolic Syndrome* cluster variants colocalised in adipose subcutaneous (*JAZF1*, *ALKAL2*, *LCORL*). The *Higher adiposity* cluster shows colocalization in skin (*MYO19*, *EIF3C*, *SLC39A10*) and whole blood (*WFS1*, *CCDC134*, *MED27*). The *Vascular dysfunction* cluster colocalised with fibroblasts (*ERI1*, *FOXD4*, *RSRC1*) and thyroid (*CSTB*, *ZNF638*, *SNX31*) and the *Reduced beta-cell function* cluster with pancreatic islets (*C2CD4B*, *ADCY5*, *PHB*).

We identified 99 tissue-specific colocalizations (***Figure 3a*** bottom, ***Methods***). While thyroid and adipose subcutaneous tissues showed a high number of total colocalizations, pancreatic islets showed the highest number (14) of single-tissue (*i.e.*, specific) colocalizations, particularly among the clusters strongly associated with risk of T2D such as the *Reduced beta-cell function*, *Metabolic Syndrome* and *Higher adiposity* clusters. Notably, the pancreatic islet tissue-specific colocalised genes include *TH* (synthesis of catecholamines)^46^ in the *Higher adiposity* cluster, *MTNR1B* (circadian rhythms and glucose metabolism)^47^, *FXYD2* (Na,K-ATPase pump regulator)^48^, *G3BP2* (cellular stress)^49^ in the *Reduced beta-cell function* cluster, *SYNDIG1L* (synapse development), *LTBP3* (cell growth, differentiation and repair)^50^, *CLEC18A* (immune function)^51^ in the *Metabolic Syndrome* cluster (***Supplementary Table 8***). This demonstrates the predominant role of pancreatic islets in T2D pathogenesis and its related complications.

To explore the underlying mechanisms in the *Inverse T2D-BP risk* cluster, we conducted pathway analysis using *Metascape*^52^ for the 202 colocalised genes identified within this cluster (***Supplementary Figure 7***). This analysis revealed an overwhelming enrichment in the retinol metabolic process (GO:0042572), which involves one of three compounds that make up vitamin A (retinol, retinal, and retinoic acid). All components of the retinol metabolism are associated with both T2D and CVD^53^.

We then dissected the localisation of our SNVs in a cluster-specific manner using chromatin accessibility atlases from CATLAS, based on scATAC-seq peaks. The atlas encompasses 222 cell types from 30 human adult tissues and 15 fetal tissues, allowing to examine the enrichment of candidate cis-regulatory elements (cCREs) in each cluster across different cell types (***Figure 3b, Supplementary Table 9***). The clusters were enriched in diverse regulatory mechanisms. Specifically, the *Inverse T2D-BP* cluster exhibited significant (P-value ≤ 2.25×10^-4^) enrichment for regions of open chromatin in mesothelial cells, endocardial cells, endothelial in exocrine tissue cells as well as fetal endocardial and mesangial cells. The *Reduced beta-cell function* cluster demonstrated strong enrichment in several fetal cell types, such as islets, gastric goblet, alveolar epithelial, cardiomyocyte, as well as follicular cells and cells from the pancreas tissues, including delta, gamma, beta and alpha. This suggests that, in addition to the islet dysregulation pathways of insulin impairment, multiple fetal development pathways are important for adult metabolic health. Moreover, nominal enrichments were observed in fetal adrenal cortical cells for the *Metabolic Syndrome* and *Higher adiposity* clusters, suggesting hormone regulatory implications beginning as early as intrauterine development^54^.

Given the large number of colocalised SNVs observed across clusters in tissues such as the thyroid, subcutaneous adipose tissue, tibial artery, tibial nerve, and lower leg skin (***Figure 3a***), we further explored the colocalised genes by identifying their enrichment in the primary cell types corresponding to these tissues – namely, follicular cells, adipocytes, smooth muscle cells, Schwann cells, keratinocytes (***Figure 3b***). Notably, we identified 15 genes that both colocalised in thyroid and were enriched in follicular cell cCREs (***Supplementary Table 10***). While some of these genes were previously associated with T2D such as *CAMK1D* (energy homeostasis and beta-cell receptor signalling pathway)^55^ or *KCNH6* (insulin secretion and glucose homeostasis)^56^, and with BP regulation such as *ACE* (renin-angiotensin system)^28^, the remaining are new potential candidate genes for the T2D-BP pathogenesis. Among them are *SAE1* (known in cancer)^57^, *GSAP* (known in Alzheimer’s disease)^58^, *DCAF7* (cellular differentiation)^59^, *MAP3K3* (stress and inflammation)^60^. Subsequent *Metascape*^52^ pathway analysis highlighted fundamental cellular processes, including protein ubiquitination (GO:0016567) and positive regulation of protein modification process (GO:0031401). The overlap of signals between colocalised genes and cCREs in the other four tissues consistently highlighted the presence of the *MAP3K3* gene. Subsequent pathway analyses did not yield conclusive results (***Supplementary Table 11***).

Overall, changes in gene expression within clusters highlighted the importance of the thyroid tissue in T2D-BP shared pathophysiology and retinol metabolism within the *Inverse T2D-BP risk* cluster, while the high number of fetal cell regulatory elements enrichment suggests a strong contribution of intrauterine growth pathways in both T2D and high BP.

### T2D-BP comorbidity using partitioned PGSs

To evaluate the ability of the T2D-BP SNV clusters to predict comorbidities and complications, we used the individual-level data from UKB and built partitioned PGSs for each of cluster group. Using the R software environment tool, *comorbidPGS*, we set unweighted partitioned PGS aligned to the T2D risk allele, *i.e.*, each allele increasing the risk of T2D is counted as one in the PGS calculation (***Methods***)^24^.

To illustrate the influence of genetic predisposition, we computed the relative risk of comorbidity among the UKB individuals (***Methods***), based on the top 10% percentiles of each partitioned PGS. Whereas the overall prevalence of T2D-high BP comorbidity in the UKB was 5.49% (***Figure 4a***), individuals in top ten percent of the unweighted risk score of *Higher adiposity*, *Metabolic Syndrome*, and *Reduced beta-cell function* clusters had a relative risk RR[95%CI] of 1.36[1.32-1.41], 1.44[1.39-1.48], 1.55[1.51-1.60], respectively. Moreover, the individuals in the top 10% distribution of *Metabolic Syndrome* and *Reduced beta-cells function* combined PGSs, derived from 536 SNVs, showed a 2.13[1.96-2.31] fold increased risk of having comorbidity (***Figure 4b***). Survival analysis, using cumulative hazard plots, indicated that this elevated comorbidity risk was consistent and linear over 15 years of follow-up (year 0 representing the date of first diagnosis with either hypertension or T2D, ***Figure 4c***). This suggests that individuals with high PGS distributions remain at increased risk of comorbidity throughout their life course. Consequently, partitioned PGSs could have clinical relevance in identifying younger individuals at high risk more effectively than existing methods^61,62^.

**Figure 4.**
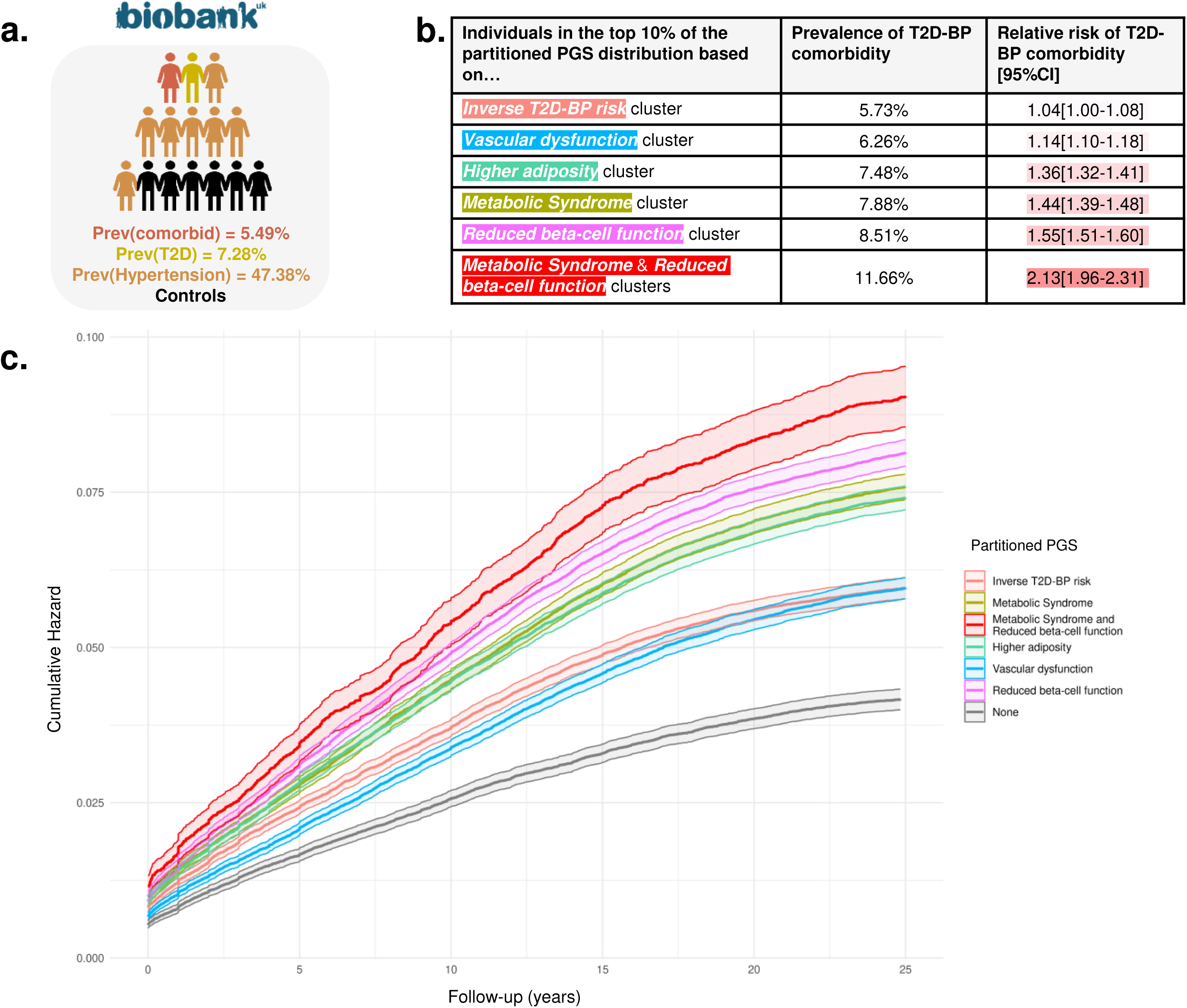
The T2D-BP comorbidity risks stratified by partitioned PGSs in the UKB. **a**, Overall prevalence of individuals with T2D, hypertension, and T2D-BP comorbidity. Prevalence (Prev) describes the proportion of cases divided by the total, 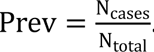. **b,** Summary table depicting the risk of T2D-BP comorbidity based on being in the top ten percent of cluster PGSs in the UKB. Relative risk (RR) is calculated as the ratio of the proportion of cases within a cluster subgroup to the proportion of cases in the overall population, RR = 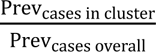. **c**, Cumulative hazard plot of T2D-BP comorbidity stratified by being in the top 33 percent of the unweighted partitioned PGS after clustering. <u>Legend:</u> Prev = prevalence; comorbid = T2D-BP comorbidity; T2D = type 2 diabetes; BP = blood pressure; CI = confidence interval.

Using the partitioned PGSs, we evaluated the association between PGS and multiple sets of complications based on the UKB hospital records (***Figure 5***, ***Supplementary Table 12***). We detected a reciprocal protective effect of the ***Inverse T2D-BP* cluster** PGS on essential hypertension (OR[95% CI]=0.91[0.90-0.92], P<1.00×10^-40^) alongside other circulatory system disorders such as coronary artery disease (CAD, OR[95% CI]=0.94[0.93-0.95], P=5.08×10^-22^), angina pectoris (OR[95% CI]=0.95[0.94-0.97], P=1.29×10^-12^), AF (OR[95% CI]=0.96[0.94-0.97], P=1.01×10^-10^), chronic ischemic heart disease (OR[95% CI]=0.96[0.95-0.97], P=9.08×10^-10^). The *Inverse T2D-BP risk* cluster PGS also associates with lower risk of gout (OR[95% CI]=0.92[0.89-0.95], P=1.10×10^-6^) and hypercholesterolaemia (OR[95% CI]=0.98[0.97-0.99], P=1.90×10^-17^). These results support previous research on the heterogeneous effects of hypertensive medications on the risk of T2D, indicating that some biological processes between T2D and high BP may reduce the risk of comorbidity^16^.

**Figure 5.**
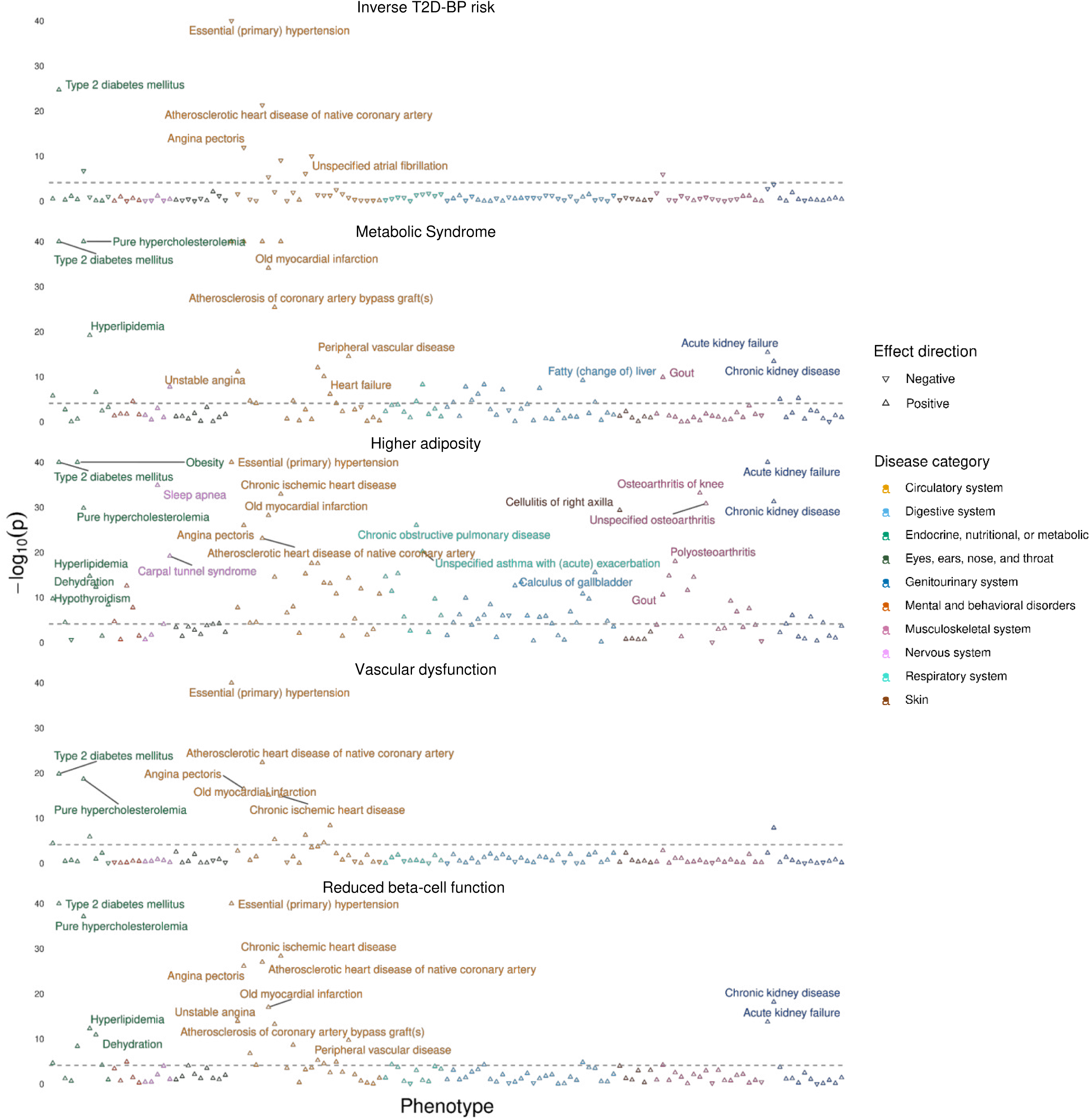
Association between complications and partitioned PGSs after clustering in the UKB. The figure displays five Manhattan plots, each representing one of the five clusters. The grey dotted line indicates the threshold for significant associations. Arrows illustrate the direction of association between PGS and disease risk: an upward-pointing arrow signifies that a higher PGS is associated with an increased risk of the disease, while a downward-pointing arrow indicates that a higher PGS is associated with a decreased risk of the disease. The colours differentiate between the different disease categories. <u>Legend:</u> PGS = polygenic score; UKB = UK Biobank; T2D = type 2 diabetes; BP = blood pressure.

We confirmed the high contribution into T2D-BP comorbidity of the ***Metabolic Syndrome* cluster**, by showing that its relatively small number of SNVs could predict a higher risk of multiple metabolic disorders, including T2D (OR[95% CI]=1.24[1.23-1.26], P<1.00×10^-40^), hypertension (OR[95% CI]=1.13[1.12-1.14], P≤1.00×10^-40^), hypercholesterolaemia (OR[95% CI]=1.10[1.09-1.10], P≤1.00×10^-40^), hyperlipidaemia (OR[95% CI]=1.10[1.08-1.13], P=6.52×10^-20^), fatty liver (OR[95% CI]=1.13[1.09-1.18], P=6.08×10^-10^), and hypothyroidism (OR[95% CI]=1.03[1.02-1.04], P=1.65×10^-6^). Additionally, the *Metabolic Syndrome* cluster PGS showed significant association with higher risk of cardiovascular complications such as CAD, angina pectoris, ischemic heart disease, heart failure, and myocardial infarction. We also detected association with higher risk of kidney failure and calculus of kidney.

The ***Higher adiposity* PGS** showed the strongest risk prediction of obesity-related diseases, including T2D (OR[95% CI]=1.22[1.20-1.23], P≤1.00×10^-40^), hypertension (OR[95% CI]=1.09[1.09-1.10], P≤1.00×10^-40^), sleep apnea (OR[95% CI]=1.17[1.14-1.20], P=1.28×10^-34^), osteoarthritis (OR[95% CI]=1.08[1.07-1.10], P=1.51×10^-31^), carpal tunnel syndrome (OR[95% CI]=1.09[1.07-1.11], P=6.61×10^-20^), and pneumonia (OR[95% CI]=1.08[1.06-1.10], P=3.92×10^-12^). This cluster PGS was associated with higher mental disorders, such as major depressive disorder, delirium or behavioural disorders due to use of tobacco, highlighting the intertwined relations between obesity and depressive conditions.

Among other clusters, the ***Vascular Dysfunction* cluster** was more predictive for cardiovascular complications, including hypertension (OR[95% CI]=1.12[1.11-1.13], P<1.00×10^-40^), CAD (OR[95% CI]=1.06[1.05-1.07], P=4.25×10^-23^), or ischemic heart disease (OR[95% CI]=1.06[1.04-1.07], P=1.05×10^-15^). The ***Reduced beta-cell function* unweighted PGS** showed the strongest association with risk of T2D (OR[95% CI]=1.33[1.32-1.35], P<1.00×10^-40^) and its related consequences, including obesity (OR[95% CI]=1.03[1.02-1.05], P=4.38×10^-9^), hyperlipidaemia (OR[95% CI]=1.08[1.06-1.10], P=5.17×10^-13^), fatty liver (OR[95% CI]=1.09[1.05-1.14], P=1.62×10^-5^), chronic kidney disease (OR[95% CI]=1.10[1.07-1.12], P=6.47×10^-19^), and hypothyroidism (OR[95% CI]=1.03[1.01-1.04], P=2.43×10^-5^). The *Reduced beta-cell function* weighted PGS (***Methods***) showed strong association with high SBP and PP, albeit not with DBP (***Supplementary Table 13***).

The unweighted PGS effectively delineated the differences among the T2D-BP cluster SNVs. The partitioning of PGSs in T2D-BP comorbidity highlighted that a reduced number of SNVs can predict multiple related conditions at young age through diverse pathophysiological processes.

## Discussion

In this large-scale multiomic study, we explored the complex genetic underpinnings of the comorbid relationship between T2D and high BP. To our knowledge, this is the first large-scale genetic analysis to dissect the shared genetic architecture of these two closely related metabolic conditions. Our findings corroborate previous studies on genetic correlation, revealing a robust genetic correlation and large overlap in genetic signals between T2D and high BP^63,64^.

We curated a set of SNVs associated with T2D and high BP. By using this refined SNV list, we prioritised common variants with stronger associations and mitigate the pleiotropy effects. Through hierarchical clustering using T2D-BP related endpoints and risk factors, we identified five clusters of SNVs, each highlighting unique pathogenetic processes underlying the T2D-BP relationship. This clustering approach provided a clearer delineation of genetic relationships, reducing heterogeneity compared to other clustering methods. Four of these SNV clusters – *Metabolic Syndrome*, *Higher adiposity*, *Vascular dysfunction*, and *Reduced beta-cell function* – align with established findings in high BP or T2D^27,65–67^. We discovered an intriguing cluster of variants with an *Inverse T2D-BP risk*, which suggest a potential perturbation in retinol metabolism. Although recent meta-analysis on the role of retinol role in T2D and BP regulation highlighted inconsistent results, one retinol derivative, retinoic acid, has been consistently linked to higher IR and reduced cardiovascular events^53^. The *Metabolic Syndrome* cluster was associated with novel features such as shorter stature, higher WHR, and no detectable effect on BMI. Additionally, we observed an enrichment in colocalizations specific to the thyroid across all five T2D-BP clusters, suggesting a potential mechanistic role for thyroid function in T2D-BP comorbidity. This feature further highlights a need for better thyroid health in general population to reduce impact of dysthyroidism on BP and T2D management^68^.

We bring forward a property of partitioned PGS to differentially predict related T2D – high BP conditions, highlighting their potential utility in risk prediction at a young age^69^. While PGS has shown predictive value for an expanding array of common diseases, such as for instance CAD^70,71^, its clinical application remains limited due to a range of pitfalls^72^. In this study, we report how specific clusters, particularly the *Metabolic Syndrome* and *Reduced beta-cell function* clusters, more effectively identify a sub-population of individuals over twice the general population risk of T2D-BP comorbidity. Our study calls for partitioning of PGSs to predict complications in metabolic disorders like T2D and high BP, suggesting that in comorbidity risks, less SNVs are more impactful than an entire genome-wide PGS to stratify the individuals at high risk of comorbidity. This work paves the way for improved risk stratification and precision health approaches^71^.

We acknowledge several limitations in this work. The metabolic mechanisms involved in both T2D and high BP are not fully explained by genetics alone, and are influenced by environmental factors such as salt consumption or western diet. Moreover, the heterogeneity in individual origins and sample sizes could potentially reduce our ability to identify clear pathways. To address this concern, we consistently incorporated GWAS summary statistics with multiple ancestries including individuals of European, East Asian, South Asian, and African descent^73^. Hierarchical clustering can ‘force’ an SNV into fit a cluster that may not fully capture its effect. To mitigate this, we compared and found consistent results with previous clustering studies and sensitivity analyses using other clustering methods, such as bNMF^27^.

In this study, we highlight the mechanistic heterogeneity in T2D and high BP, identifying five distinct genetic pathways that contribute to the risk of T2D-BP comorbidity, each associated with specific physiological alterations. The partitioned PGSs, based on clustering, demonstrate improved accuracy in estimating lifetime risk trajectories for T2D-BP comorbidity. These findings provide a foundation for precision medicine approaches tailored to the unique pathophysiology of each pathway.

## Online Methods

### Material description

We collected 49 publicly available GWAS summary statistics (***Supplementary Table 4***), spanning from 2019 to 09 June 2023. GWAS were selected if they included a large number of participants (N ≥ 10,000) and a preference for datasets with diverse ancestral backgrounds to enhance genetic diversity in our study, primarily involving individuals of European, East Asian, South Asian, and African descent. European-only studies were included when multi-ancestry data were unavailable.

To ensure robustness in uncovering shared aetiologies, we curated from multiple sources a list of T2D and BP SNVs reaching genome-wide significance (*i.e.*, P-value < 5×10^-8^) and characterised by prevalence (minor allele frequency (MAF) > 0.01). We identified a total of 1,401 SNVs. This list was further curated for clustering to keep only independent SNVs (LD r² < 0.2). The independent SNV list encompasses 1,304 SNVs, including 500 T2D and 283/272/270 SBP/DBP/PP signals, with 9 unique SNVs demonstrating associations with T2D-BP comorbidity.

The UK Biobank (UKB, https://ukbiobank.ac.uk/) is a large prospective cohort study with genotypic and phenotypic data^21^. The dataset in this study derived from genome-wide imputed data, including 459,247 individuals of European ancestry. To identify individuals with T2D, we leveraged hospital admission records, self-reports, and ICD10/9 codes, successfully defining 33,446 T2D individuals. Hypertension was assessed using three blood pressure metrics: SBP, DBP, and PP, with both automated and manual records available in the UKB^12^. We used the mean value when more than one value was found for a given individual and adjusted for medication use by adding 15 mmHg to SBP and 10 mmHg to DBP for individuals with reported data on blood pressure-lowering medication^74^. We identified 217,599 hypertensive individuals (***Supplementary Table 1***). The exact criteria used to identify T2D and hypertensive cases are depicted in ***Supplementary Figure 8***.

The Genotype-Tissue Expression (GTEx, https://www.gtexportal.org/home/datasets/) is a project gathering samples from 49 non-diseased tissue across 1,000 deceased individuals^17^. We used the expression quantitative trait *loci* (eQTLs) mapping genetic variants with changes in the expression of nearby genes.

The ABOS cohort is an ongoing prospective study that aims to identify the determinants of bariatric surgery outcomes, initiated at Lille University Hospital (Lille, France) in 2006. The study protocol has been previously detailed elsewhere (clinicaltrials.gov, NCT01129297)^19^. 372 individuals of European descent were included in the ABOS liver eQTL study.

The translational human pancreatic islet genotype tissue-expression resource (TIGER, http://tiger.bsc.es/) is a large meta-analysis of cohorts aggregating more than 500 human islet genomic datasets from five cohorts in the Horizon 2020 consortium T2DSystems^18^.

The cis-element ATLAS (CATLAS, http://catlas.org/humanenhancer/) is a comprehensive resource gathering the genome-wide cCREs in 222 human cell types. The dataset encompasses chromatin accessibility data derived from single-cell Assay for Transposase-Accessible Chromatin (ATAC-seq) peaks spanning 30 human adult and 15 human fetal tissues.

We systematically applied multiple correction testing and adjusted the appropriate individual-level associations for age, sex, genotyping array and the first six principal components derived from genetic data.

### Genetic correlation

We conducted genetic correlation analysis by using the whole T2D^22^ and BP^12^ GWAS datasets of European ancestry. We estimated the liability-scale heritability (ℎ_2_) of each phenotype and their genetic correlation (*r*_*g*_) by employing linkage disequilibrium score regression via *ldsc* v1.0.1^23^. We used pre-computed LD scores from 1000 Genome Phase 3 SNVs in individuals of European ancestry. We manually aligned the GWAS datasets and excluded outlier SNVs, that are multi-allelic, poorly-imputed (by filtering to HapMap3 SNVs), have a MAF > 0.01 and 0 < P-value ≤ 1.

### Reciprocal risk prediction using PGS

We constructed PGS before and after clustering with *plink* v1.9^75^, using specific SNV sets, weights from T2D^22^, SBP, DBP, and PP^67^ summary statistics and individual-level data from the UKB. We carefully selected weights without UKB to avoid sample overlap between the base data (composed of GWAS significant SNVs plus their weights) and the target data (individual-level data from the UKB). The Pruning and Thresholding (P+T) method was used before making PGS by selecting the only GWAS significant SNVs for each desired outcome^76,77^. We subsequently used *comorbidPGS* to conduct linear or logistic regression to evaluate the shared predisposition between PGS for the i-th trait and the j-th target phenotype Y, correcting for covariates including age, gender, genetic array, and the first six principal components as following^24^:

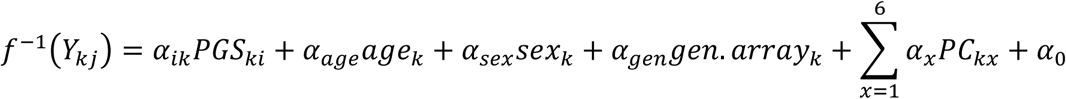

Where *Y*_*kj*_ is the value of the j-th phenotype of the k-th individual (1 or 0 for T2D, continuous value in mmHg for BP traits), *PGS_ki_* is the PGS for the i-th trait and the k-th individual, *age_k_*, *sex_k_*, *gen. array*_*k*_ and *PC*_*kx*_ being the respective covariates for the k-th individual. The α are the regression coefficients, α_0_ is the intercept.

### Genetic overlap

We estimated the genetic overlap among the pre-curated non-independent 1,401 SNVs by identifying variants within 500 kb of each other, or in LD r²>0.2 for individuals of European ancestry. The reported *loci* are the proximal ones, identified by *biomaRt*^78^. We identified and coloured the signals per clusters in ***Supplementary Table 3***.

### Clusters of pathogenetic processes

We aggregated the 1,304 independent T2D-BP SNVs and investigated 49 endophenotypes GWAS summary statistics to cluster them into distinct groups based on pathogenetic processes. Initially, we harmonised the SNVs by finding strong proxies (LD r² > 0.6) prevalent across all or most of the GWASs. For the i-th SNV of the j-th trait, we extracted the beta coefficient, β_*ij*_, along with its standard error, σ_*ij*_, to build z-scores using the following formula 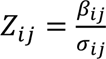. Any signal exhibiting more than 20% missingness across the GWAS was excluded. We conducted imputation of the remaining missing z-scores using a random forest algorithm from *imputeSCOPA*^34^. For each GWAS, we truncated SNVs if their absolute z-score exceeded two standard deviations.

We performed hierarchical clustering with the Ward method from the R function *hclust*, using Euclidian distance and the R package *pheatmap* for plotting (***Supplementary Figure 1***). We determined the number of clusters by taking half of the maximum Euclidian tree-row distance. Our methodology, termed ‘hard clustering’, ensures precise assignment of each SNV to a single cluster. Each variant is treated as a unit, unlike approaches that treat positive and negative associations independently. Using a ‘hard clustering’ method offers a clearer delineation of clustering patterns. However, it may force a SNV into a group where is does not truly belong^9,37^. To validate our clusters, we compared them with those identified in the latest T2DGGI study^9^ and latest T2D ‘soft’ clustering^37^. We performed GWAS weight comparison between our T2D-BP clusters and the T2DGGI clusters, and SNV assignment comparison between our T2D-BP clusters, T2DGGI cluster, and latest T2D ‘soft’ clusters. For GWAS weight comparison, we extracted each cluster weights for the common GWAS endophenotypes used in the T2DGGI clustering (k=8) and ours (k=5). We then calculated the Pearson correlation coefficient between the sets of trait cluster weights. Notably, our T2D-specific clusters showed significant correlations: the *Higher adiposity* cluster aligned with T2DGGI ‘Obesity’ cluster, the *Metabolic Syndrome* cluster was correlated with ‘Lipodystrophy’, ‘Metabolic syndrome’, and ‘Residual glycaemic’. The *Reduced beta-cell* cluster exhibited correlation across all T2DGGI clusters, particularly with ‘Residual glycaemic’ and ‘Obesity’ (***Supplementary Figure 2***). For SNV assignment comparison, we systematically looked for LD proxies (LD r²>0.6) in both T2DGGI and ‘soft’ clusters. We assign a SNV into ‘soft’ clusters based on weight > 0.75 (***Supplementary Table 5***, ***Supplementary Figure 3***).

Sensitivity analyses encompass SNV pruning by taking only the ones with association for BP, T2D or both (absolute z-score ≥ 1.645), and using two alternative clustering methods: *MRClust*^79^ (***Supplementary Figure 3***) and a ‘soft clustering’ method, bNMF^27^ (***Supplementary Figure 4***). We compared bNMF results (k=8) with hierarchical clusters (k=5) by merging the variant weights for each cluster of both methods. Logistic regression models were then employed to assess the association between the ‘hard’ cluster memberships and the ‘soft’ clustering weights. Specifically, for each hierarchical cluster, we modelled the binary cluster membership as a function of the corresponding bNMF-derived weights using a logistic regression framework. The models’ deviances were compared through ANOVA to evaluate the enrichment and significance of the clustering concordance (***Supplementary Figure 4.c***).

To improve interpretability of the clustering, we performed linear regression with the *lm* R function, across the SNVs within the k-th cluster with the j-th phenotype, defined by *E*(*Z*_*ij*_) = ∑_*k*_ α_*jk*_ *C*_*ki*_, where *C*_*ki*_ an variable taking the value 1 if the i-th SNV was assigned to the k-th cluster, 0 otherwise. The resulting regression is represented in ***Figure 2*** heatmap, with the P-value denoting the colour intensity, and the colour indicating the sign of the α_*jk*_ coefficient.

### Colocalization analysis

To identify the signals that share the same causal variants in T2D-BP GWASs and eQTLs, we systematically conducted Bayesian colocalization with *coloc.abf*^45^, between the clustered SNVs and eQTLs derived from 50 tissues from the GTEx (48 tissues), ABOS (liver) and TIGER (pancreatic islet) datasets, using a hypothesis-free approach. We assessed colocalization with the ‘lead’ GWAS per cluster (*i.e.*, the main origin of the clustered SNVs, SBP GWAS for *Inverse T2D-BP* and *Vascular dysfunction*, T2D GWAS for *Metabolic syndrome*, *Higher adiposity* and *Beta cell* clusters). For each SNV within the 1,304 clustered SNVs, we considered a genomic region of 100 kb up and downstream, and evaluated whether the variant colocalised with genetic expression in that region. Colocalization was asserted if the region was well-characterised (containing between 100 and 1,000 SNV within eQTL), the posterior probability of the model sharing a single causal variant (H4) exceeded 80%, and the posterior probability of distinct causal variants with both traits (H3) was below 50%. We excluded variants within the MHC region (chr6: 25,000,000–35,000,000).

We present in ***Figure 3a*** the count of colocalised genes across tissues (bar) and clusters (colours). We represented in this figure the top tissues based on their overall number of colocalization (N_coloc_≥100). The lower portion depicts the number of tissue-specific colocalizations, *i.e.*, when the index SNV is associated to only one tissue-specific eQTL. The complete number of colocalization per cluster and tissue is represented in ***Supplementary*** Figure 5. The colour intensity indicates the percentage of colocalised genes per cluster. Caution is warranted in interpreting the results as there is currently no robust method to rule out horizontal pleiotropy^80^.

Subsequent pathway analysis of the thyroid eQTL and *Inverse T2D-BP risk* cluster colocalised genes was performed using *metascape*^52^. For thyroid eQTL, we identified 244 colocalised genes and 75 SNVs in cCRE regions in follicular cells (used as major cell type within the thyroid). Among them, 15 signals were both associated with a colocalised gene in thyroid and a cCRE region in follicular cells. We used these remaining 15 colocalised genes with *metascape* default parameters. We performed the pathway analysis using 926 background genes expressed in the GTEx Thyroid RNA-seq v8 (TPM>120). Similar analyses were conducted for the four other top colocalised tissues: adipose subcutaneous (adipocytes as major cell type), artery tibial (smooth muscle cells), nerve tibial (Schwann general cells), and skin (sun-exposed) lower leg (keratinocytes as major cell type). Pathway analyses were performed using *metascape*, with background genes expressed in GTEx RNA-seq v8 for each respective tissue: 1,053 genes for adipose subcutaneous, 1,053 for artery tibial, 1,029 for nerve tibial, and 791 genes for skin (TPM>120).

### scATAC-seq enrichment analysis

To gain insights in the associated regulatory processes, we took the 1,304 clustered SNVs to look for enrichment of regions of open chromatin within clusters. Specifically, we aggregated independent SNVs (LD r²<0.2) within 50 kb of each clustered SNV from the 1000 Genome Project Phase 3. We followed a similar protocol by the T2DGGI paper^9^, and conducted a Firth bias-reduced logistic regression, using *logistf* R package and the following equation:

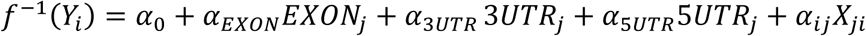

With *Y*_*j*_ taking the value 1 if the i-th SNV is within one of the clusters, *X*_*ji*_ taking the value 1 if the i-th SNV mapped to an ATAC-seq peak for the i-th cell type, 0 otherwise. We defined *EXON*_*j*_, 3*UTR*_*j*_, 5*UTR*_*j*_ indicators taking the value 1 if the j-th SNV is located to the respective annotation (from the Ensembl Project), 0 otherwise. The α are the coefficients of log fold enrichments, and α_0_ is the intercept.

We performed the logistic regression twice, one with α_*ij*_ = 0 and one without constraint. We reported on the heatmap of ***Figure 3b*** the P-value associated to Chi-squared tests between the two logistic regression models to observe the enrichment of the i-th cell type across clusters. We used multiple correction testing for Chi-squared tests across cell types and clusters, P-value_threshold_ = 2.25×10^-4^. More information on the 222 cell types gathered in CATLAS can be found in <u>CATLAS</u> website.

### Prevalence and relative risk of T2D-BP comorbidity using partitioned PGS

The prevalence of T2D-BP comorbidity was defined by the proportion of cases divided by the total number of individuals in the UKB, 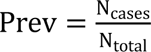. The summary table in ***Figure 4b*** provides the prevalence of individuals with T2D-BP comorbidity (of having both T2D and hypertension) based on being in the top decile (top 10%) of the partitioned PGS distribution. For individuals associated with multiple clusters, assignment was based on the cluster where they ranked the highest. The relative risk (RR) is calculated as the ratio of the proportion of cases within a cluster subgroup to the proportion of cases in the overall population 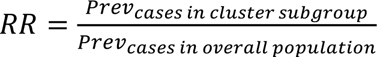. The 95% confidence interval of the RR was estimated using the following equation:

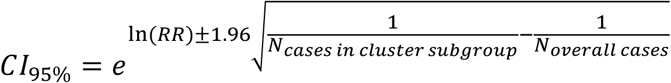

### Survival analysis using partitioned PGS

Survival analysis was conducted using the *survival* R package v3.6-4 with a Cox proportional hazards model. In this model, time zero was defined as the date of the first diagnosis of either hypertension or type 2 diabetes (T2D). The primary event of interest was the subsequent development of a T2D-BP comorbidity, marked by the diagnosis of the second disease. To ensure adequate sample sizes within groups, individuals were assigned to a cluster group if their maximum partitioned PGSs was in the top 33% of distribution. For individuals associated with multiple clusters, assignment was based on the cluster where they ranked the highest. ***Figure 4c*** displays the cumulative hazard with 95% confidence intervals.

### Partitioned PGS and risk of complications

Following the same pipeline developed in ***Reciprocal risk prediction using PGS***, partitioned PGSs were calculated using clustered SNVs in the risk-increasing direction of T2D, assuming an additive model (***Supplementary Table 12***). We do not associate a weight with partitioned PGSs, meaning each allele increasing the risk of T2D is counting as one. Sensitivity analysis included using weights derived from T2D and BP GWAS to predict the risk of the other disorders (***Supplementary Table 13***). Association with complications were performed using the UKB ICD-10 codes, updated with hospital data up to December 2023. Associations were corrected for covariates including age, gender, genetic array, and the first six principal components. Briefly, we extracted all the UKB ICD-10 available codes and refined them to subcategories of interest, namely: Endocrine, nutritional, or metabolic; Mental and behavioural disorders; Nervous system, Eyes, ears, nose and throat; Circulatory system; Respiratory system; Digestive system; Skin; Musculoskeletal system; Genitourinary system. All reported associations in ***Figure 5*** were derived using unweighted partitioned PGS, *comorbidPGS* using binary logistic regression, and a P-value corrected for multiple testing, P-value_threshold_= 7.75×10^-5^.

## Statements

## Acknowledgement

This research has been conducted using the UK Biobank Resource under application number 236. This project was in part funded by the Agence Nationale de la Recherche under the Programme d’Investissement d’Avenir (PreciDIAB, ANR-18-IBHU-0001 and RHU PreciNASH ANR-16-RHUS-0006), by the European Union through the “Fonds Européen de Développement Regional” (FEDER), by the “Conseil Régional des Hauts-de-France” (Hauts-de-France Regional Council), by the “Métropole Européenne de Lille” (MEL, European Metropolis of Lille), and by the European Research Council (ERC OpiO – 101043671, to AB)

The authors would like to thank all the investigators from different consortia that built and shared the GWAS meta-analysis, eQTLs, and scATAC-seq atlases used in this study, as well as the UK Biobank participants and dedicated staff.

## Statement of Ethics

A Statement of Ethics is not applicable because this study is based exclusively on published literature.

## Data availability

The GWAS used in this study are all publicly available and listed in ***Supplementary Table 4***. The UK Biobank Resource (UKB, https://ukbiobank.ac.uk/) was accessed using the Application Number 236. GTEx (https://www.gtexportal.org/home/datasets/) and TIGER (http://tiger.bsc.es/) eQTLs are publicly available. Data from the ABOS cohort are not publicly available, as the study is ongoing. The ATAC-seq data from CATLAS are publicly available http://catlas.org/humanenhancer/.

## Code availability

The main code used in this analysis can be found in this GitHub repository: https://github.com/VP-biostat/T2D-BP-Manuscript.

## Author Contributions

V Pascat designed the experiments and wrote the paper. L Zudina contributed to the study design for clustering and partitioned PGS. L Maurin contributed to the analysis for colocalization, pathway analysis, interpretation, and revision. A Ulrich, J Maina, A Demirkan, Z Balkhiyarova, I Pupko, and Y Sharhorodska defined the phenotypes of interest in the UK Biobank and provided handmade GWAS summary statistics. F Pattou and B Staels provided the ABOS cohort. M Kaakinen contributed to the overall statistics evaluation. A Abdelgader-Khamis contributed to the colocalization evaluation. A Bonnefond, P Munroe, P Froguel, and I Prokopenko contributed to evaluation of the results. V Pascat and I Prokopenko led the manuscript writing. P Froguel and I Prokopenko jointly supervised the study. All authors read and approved the final paper.

## Reference

1. the World Health Organization. Hypertension. https://www.who.int/news-room/fact-sheets/detail/hypertension (2023).

2. International Diabetes Federation. IDF Diabetes Atlas. https://diabetesatlas.org/ (2021).

3. Iglay, K. et al. Prevalence and co-prevalence of comorbidities among patients with type 2 diabetes mellitus. Curr Med Res Opin 32, 1243–1252 (2016).

4. Colussi, G. L., Da Porto, A. & Cavarape, A. Hypertension and type 2 diabetes: lights and shadows about causality. Journal of Human Hypertension vol. 34 91–93 Preprint at 10.1038/s41371-019-0268-x (2020).

5. Lastra, G., Syed, S., Kurukulasuriya, L. R., Manrique, C. & Sowers, J. R. Type 2 Diabetes Mellitus and Hypertension. Endocrinol Metab Clin North Am 43, 103–122 (2014).

6. Schmieder, R. E. et al. Achievement of individualized treatment targets in patients with comorbid type-2 diabetes and hypertension: 6 months results of the DIALOGUE registry. BMC Endocr Disord 15, 23 (2015).

7. Alberti, K. G. M. M., Zimmet, P. & Shaw, J. Metabolic syndrome - A new world-wide definition. A consensus statement from the International Diabetes Federation. Diabetic Medicine 23, 469–480 (2006).

8. Huang, P. L. A comprehensive definition for metabolic syndrome. DMM Disease Models and Mechanisms 2, 231–237 (2009).

9. Suzuki, K. et al. Genetic drivers of heterogeneity in type 2 diabetes pathophysiology. Nature 627, 347–357 (2024).

10. Keaton, J. M. et al. Genome-wide analysis in over 1 million individuals of European ancestry yields improved polygenic risk scores for blood pressure traits. Nat Genet 56, 778–791 (2024).

11. Qi, Q. et al. Genetic Predisposition to High Blood Pressure Associates With Cardiovascular Complications Among Patients With Type 2 Diabetes. Diabetes 61, 3026– 3032 (2012).

12. Evangelou, E. et al. Genetic analysis of over 1 million people identifies 535 new loci associated with blood pressure traits. Nat Genet 50, 1412–1425 (2018).

13. Ehret, G. B. et al. Genetic variants in novel pathways influence blood pressure and cardiovascular disease risk. Nature 478, 103–109 (2011).

14. Sun, D. et al. Type 2 Diabetes and Hypertension: A Study on Bidirectional Causality. Circ Res 124, 930–937 (2019).

15. Aikens, R. C. et al. Systolic Blood Pressure and Risk of Type 2 Diabetes: A Mendelian Randomization Study. Diabetes 66, 543–550 (2017).

16. Nazarzadeh, M. et al. Blood pressure lowering and risk of new-onset type 2 diabetes: an individual participant data meta-analysis. The Lancet 398, 1803–1810 (2021).

17. Aguet, F. et al. The GTEx Consortium atlas of genetic regulatory effects across human tissues. Science (1979) 369, 1318–1330 (2020).

18. Alonso, L. et al. TIGER: The gene expression regulatory variation landscape of human pancreatic islets. Cell Rep 37, 109807 (2021).

19. Margerie, D. et al. Hepatic transcriptomic signatures of statin treatment are associated with impaired glucose homeostasis in severely obese patients. BMC Med Genomics 12, 80 (2019).

20. Zhang, K. et al. A single-cell atlas of chromatin accessibility in the human genome. Cell 184, 5985–6001.e19 (2021).

21. Sudlow, C. et al. UK Biobank: An Open Access Resource for Identifying the Causes of a Wide Range of Complex Diseases of Middle and Old Age. PLoS Med 12, e1001779 (2015).

22. Vujkovic, M. et al. Discovery of 318 new risk loci for type 2 diabetes and related vascular outcomes among 1.4 million participants in a multi-ethnic meta-analysis. 52, 680–691 (2020).

23. Bulik-Sullivan, B. et al. LD score regression distinguishes confounding from polygenicity in genome-wide association studies. Nat Genet 47, 291–295 (2015).

24. Pascat, V. et al. comorbidPGS: an R package assessing shared predisposition between Phenotypes using Polygenic Scores. Hum Hered (2024) doi:10.1159/000539325.

25. Wain, L. V et al. Genome-wide association study identifies six new loci influencing pulse pressure and mean arterial pressure. Nat Genet 43, 1005–1011 (2011).

26. Mancina, R. M. et al. The COBLL1 C allele is associated with lower serum insulin levels and lower insulin resistance in overweight and obese children. Diabetes Metab Res Rev 29, 413–416 (2013).

27. Udler, M. S. et al. Type 2 diabetes genetic loci informed by multi-trait associations point to disease mechanisms and subtypes: A soft clustering analysis. PLoS Med 15, (2018).

28. Tsai, C.-T. et al. Angiotensinogen Gene Haplotype and Hypertension. Hypertension 41, 9–15 (2003).

29. Liao, Z., Wang, Y., Qi, X. & Xiao, X. JAZF1, a relevant metabolic regulator in type 2 diabetes. Diabetes Metab Res Rev 35, (2019).

30. Johnson, A. D. et al. Association of Hypertension Drug Target Genes With Blood Pressure and Hypertension in 86 588 Individuals. Hypertension 57, 903–910 (2011).

31. Lyssenko, V. et al. Mechanisms by which common variants in the TCF7L2 gene increase risk of type 2 diabetes. Journal of Clinical Investigation 117, 2155–2163 (2007).

32. Trevaskis, J. et al. Src homology 3-domain growth factor receptor-bound 2-like (endophilin) interacting protein 1, a novel neuronal protein that regulates energy balance. Endocrinology 146, 3757–64 (2005).

33. Myers, T. A., Chanock, S. J. & Machiela, M. J. LDlinkR: An R Package for Rapidly Calculating Linkage Disequilibrium Statistics in Diverse Populations. Front Genet 11, 1– 5 (2020).

34. Mägi, R. et al. SCOPA and META-SCOPA: Software for the analysis and aggregation of genome-wide association studies of multiple correlated phenotypes. BMC Bioinformatics 18, 4–11 (2017).

35. Ward, J. H. Hierarchical Grouping to Optimize an Objective Function. J Am Stat Assoc 58, 236–244 (1963).

36. Foley, C. N., Mason, A. M., Kirk, P. D. W. & Burgess, S. MR-Clust: clustering of genetic variants in Mendelian randomization with similar causal estimates. Bioinformatics 37, 531–541 (2021).

37. Smith, K. et al. Multi-ancestry polygenic mechanisms of type 2 diabetes. Nat Med 30, 1065–1074 (2024).

38. Laaksonen, D. et al. Sex hormones, inflammation and the metabolic syndrome: a population-based study. Eur J Endocrinol 601–608 (2003) doi:10.1530/eje.0.1490601.

39. Després, J.-P. & Lemieux, I. Abdominal obesity and metabolic syndrome. Nature 444, 881–887 (2006).

40. Asao, K. et al. Short Stature and the Risk of Adiposity, Insulin Resistance, and Type 2 Diabetes in Middle Age. Diabetes Care 29, 1632–1637 (2006).

41. Stefan, N., Häring, H.-U., Hu, F. B. & Schulze, M. B. Divergent associations of height with cardiometabolic disease and cancer: epidemiology, pathophysiology, and global implications. Lancet Diabetes Endocrinol 4, 457–467 (2016).

42. Ben-Shlomo, Y. et al. An investigation of fetal, postnatal and childhood growth with insulin-like growth factor I and binding protein 3 in adulthood. Clin Endocrinol (Oxf) 59, 366–373 (2003).

43. Wittenbecher, C., Kuxhaus, O., Boeing, H., Stefan, N. & Schulze, M. B. Associations of short stature and components of height with incidence of type 2 diabetes: mediating effects of cardiometabolic risk factors. Diabetologia 62, 2211–2221 (2019).

44. Johnston, L. W. et al. Short Leg Length, a Marker of Early Childhood Deprivation, Is Associated With Metabolic Disorders Underlying Type 2 Diabetes. Diabetes Care 36, 3599–3606 (2013).

45. Giambartolomei, C. et al. Bayesian Test for Colocalisation between Pairs of Genetic Association Studies Using Summary Statistics. PLoS Genet 10, e1004383 (2014).

46. Bueno-Carrasco, M. T. et al. Structural mechanism for tyrosine hydroxylase inhibition by dopamine and reactivation by Ser40 phosphorylation. Nat Commun 13, 74 (2022).

47. Hu, C. & Jia, W. Linking *MTNR1B* Variants to Diabetes: The Role of Circadian Rhythms. Diabetes 65, 1490–1492 (2016).

48. Zhou, K. et al. FXYD2 mRNA expression represents a new independent factor that affects survival of glioma patients and predicts chemosensitivity of patients to temozolomide. BMC Neurol 21, 438 (2021).

49. Kang, W. et al. Research Progress on the Structure and Function of G3BP. Front Immunol 12, (2021).

50. Zhu, G. et al. Novel LTBP3 mutations associated with thoracic aortic aneurysms and dissections. Orphanet J Rare Dis 16, 513 (2021).

51. Chang, C.-M., Chang, W.-C. & Hsieh, S. Characterization of the genetic variation and evolutionary divergence of the CLEC18 family. J Biomed Sci 31, 53 (2024).

52. Zhou, Y. et al. Metascape provides a biologist-oriented resource for the analysis of systems-level datasets. Nat Commun 10, 1523 (2019).

53. Olsen, T. & Blomhoff, R. Retinol, Retinoic Acid, and Retinol-Binding Protein 4 are Differentially Associated with Cardiovascular Disease, Type 2 Diabetes, and Obesity: An Overview of Human Studies. Advances in Nutrition 11, 644–666 (2020).

54. Horikoshi, M. et al. New loci associated with birth weight identify genetic links between intrauterine growth and adult height and metabolism. Nat Genet 45, 76–82 (2013).

55. Vivot, K. et al. CaMK1D signalling in AgRP neurons promotes ghrelin-mediated food intake. Nat Metab 5, 1045–1058 (2023).

56. Wang, H. et al. KCNH6 channel promotes insulin exocytosis via interaction with Munc18-1 independent of electrophysiological processes. Cellular and Molecular Life Sciences 81, 86 (2024).

57. Yang, Y. et al. SAE1 promotes human glioma progression through activating AKT SUMOylation-mediated signaling pathways. Cell Communication and Signaling 17, 82 (2019).

58. He, G. et al. Gamma-secretase activating protein is a therapeutic target for Alzheimer’s disease. Nature 467, 95–98 (2010).

59. Melo-Cardenas, J., Bezavada, L., Cotton, A. & Crispino, J. D. DDB1 and CUL4 Associated Factor 7 (DCAF7) Is Essential for Hematopoiesis. Blood 140, 8586–8587 (2022).

60. Guan, J., Fan, Y., Wang, S. & Zhou, F. Functions of MAP3Ks in antiviral immunity. Immunol Res 71, 814–832 (2023).

61. Jiang, X., Holmes, C. & McVean, G. The impact of age on genetic risk for common diseases. PLoS Genet 17, e1009723 (2021).

62. Thompson, D. J. et al. UK Biobank release and systematic evaluation of optimised polygenic risk scores for 53 diseases and quantitative traits. doi:10.1101/2022.06.16.22276246.

63. Wielscher, M. et al. Genetic correlation and causal relationships between cardio-metabolic traits and lung function impairment. Genome Med 13, 104 (2021).

64. Vattikuti, S., Guo, J. & Chow, C. C. Heritability and Genetic Correlations Explained by Common SNPs for Metabolic Syndrome Traits. PLoS Genet 8, e1002637 (2012).

65. Scott, R. A. et al. An Expanded Genome-Wide Association Study of Type 2 Diabetes in Europeans. Diabetes 66, 2888–2902 (2017).

66. Dimas, A. S. et al. Impact of Type 2 Diabetes Susceptibility Variants on Quantitative Glycemic Traits Reveals Mechanistic Heterogeneity. Diabetes 63, 2158–2171 (2014).

67. Warren, H. R. et al. Genome-wide association analysis identifies novel blood pressure loci and offers biological insights into cardiovascular risk. Nat Genet 49, 403–415 (2017).

68. Gavrila, A. & Hollenberg, A. N. The Hypothalamic-Pituitary-Thyroid Axis: Physiological Regulation and Clinical Implications. in The Thyroid and Its Diseases 13–23 (Springer International Publishing, Cham, 2019). doi:10.1007/978-3-319-72102-6_2.

69. Lambert, S. A., Abraham, G. & Inouye, M. Towards clinical utility of polygenic risk scores. Hum Mol Genet 28, R133–R142 (2019).

70. Surakka, I., et al. Sex-Specific Survival Bias and Interaction Modeling in Coronary Artery Disease Risk Prediction. Circ Genom Precis Med 16, e003542 (2023).

71. Ma, Y. & Zhou, X. Genetic prediction of complex traits with polygenic scores: a statistical review. Trends in Genetics 37, 995–1011 (2021).

72. Novembre, J. et al. Addressing the challenges of polygenic scores in human genetic research. The American Journal of Human Genetics 109, 2095–2100 (2022).

73. Graham, S. E. et al. The power of genetic diversity in genome-wide association studies of lipids. Nature 600, 675–679 (2021).

74. Tobin, M. D., Sheehan, N. A., Scurrah, K. J. & Burton, P. R. Adjusting for treatment effects in studies of quantitative traits: Antihypertensive therapy and systolic blood pressure. Stat Med 24, 2911–2935 (2005).

75. Purcell, S. et al. PLINK: A tool set for whole-genome association and population-based linkage analyses. Am J Hum Genet 81, 559–575 (2007).

76. Choi, S. W. & O’Reilly, P. F. PRSice-2: Polygenic Risk Score software for biobank-scale data. Gigascience 8, 1–6 (2019).

77. Choi, S. W., Mak, T. S. H. & O’Reilly, P. F. Tutorial: a guide to performing polygenic risk score analyses. Nat Protoc 15, 2759–2772 (2020).

78. Durinck, S., Spellman, P. T., Birney, E. & Huber, W. Mapping identifiers for the integration of genomic datasets with the R/Bioconductor package biomaRt. Nat Protoc 4, 1184– 1191 (2009).

79. Foley, C. N., Mason, A. M., Kirk, P. D. W. & Burgess, S. MR-Clust: clustering of genetic variants in Mendelian randomization with similar causal estimates. Bioinformatics 37, 531–541 (2021).

80. Richardson, T. G. et al. Systematic Mendelian randomization framework elucidates hundreds of CpG sites which may mediate the influence of genetic variants on disease. Hum Mol Genet 27, 3293–3304 (2018).

