## Supplementary Figures for "Mechanistic Heterogeneity in Type 2 Diabetes and Hypertension Comorbidity Revealed with Partitioned Polygenic Scores"

### Genetic architecture of the type 2 diabetes-hypertension comorbidity

#### Supplementary Figures


**Supplementary Figure 1:** Hierarchical clustering of the 1,304 SNVs using *pheatmap*. 2

**Supplementary Figure 2:** *MRClust* results using T2D as exposure and PP as outcome using the curated 563 T2D SNVs as instrument variables (***Methods – Clusters of pathogenetic processes***). 4

**Supplementary Figure 3:** Bayesian nonnegative matrix factorization (bNMF) clustering using the 1,304 T2D-BP SNVs, ran with defaults parameters and 10 iterations (***Methods – Clusters of pathogenetic processes***). Most iterations (5/10) resulted in 8 groups. **a,** Variant association to clusters. **b,** Feature (GWAS summary statistics) association to clusters. **c,** Comparison between the hierarchical clusters (columns) and the bNMF clusters (rows). Each value intensity represents the associated -log(P-value) of the linear association while the colour is the direction of the linear association beta (***Methods – Clusters of pathogenetic processes***). 5

**Supplementary Figure 4: a,** SNV comparison between the T2D-BP hierarchical clusters (x-axis) and the cluster of the latest T2D hierarchical clustering (T2DGGI paper) from Suzuki et al. (2024). **b,** Comparison between the T2D-BP hierarchical clusters (x-axis) and the latest T2D 'soft' clustering from Smith et al. (2024). The comparison is done using each study genetic variants and looking for LD proxy (LD $r^2 > 0.6$) in our T2D-BP genetic variants. For 'soft' clusters, the cluster assignment is based on weight > 0.75 (***Methods – Clusters of pathogenetic processes***). 7

**Supplementary Figure 5:** GWAS weight comparison between the hierarchical clusters (columns) and the cluster of the T2DGGI paper (rows). Each value intensity represents the associated Pearson correlation coefficient (***Methods – Clusters of pathogenetic processes***). 8

**Supplementary Figure 6:** Heat map of colocalised loci across the five clusters and 50 human adult tissues. Each column corresponds to a cluster, while each row represents a tissue. The numerical value in each tile indicates the number of colocalised loci for a specific cluster and tissue. Colour intensity corresponds to the percentage of colocalisation per cluster. 9

**Supplementary Figure 7:** Pathway analysis of the colocalised genes within the *Inverse T2D-BP risk* cluster SBP GWAS using *metascape*. The following genes were associated with the pathway mentioned above. 10

**Supplementary Figure 8:** Diagram of criteria used in the UKB to identify T2D and hypertension cases, and adjust BP levels. 11

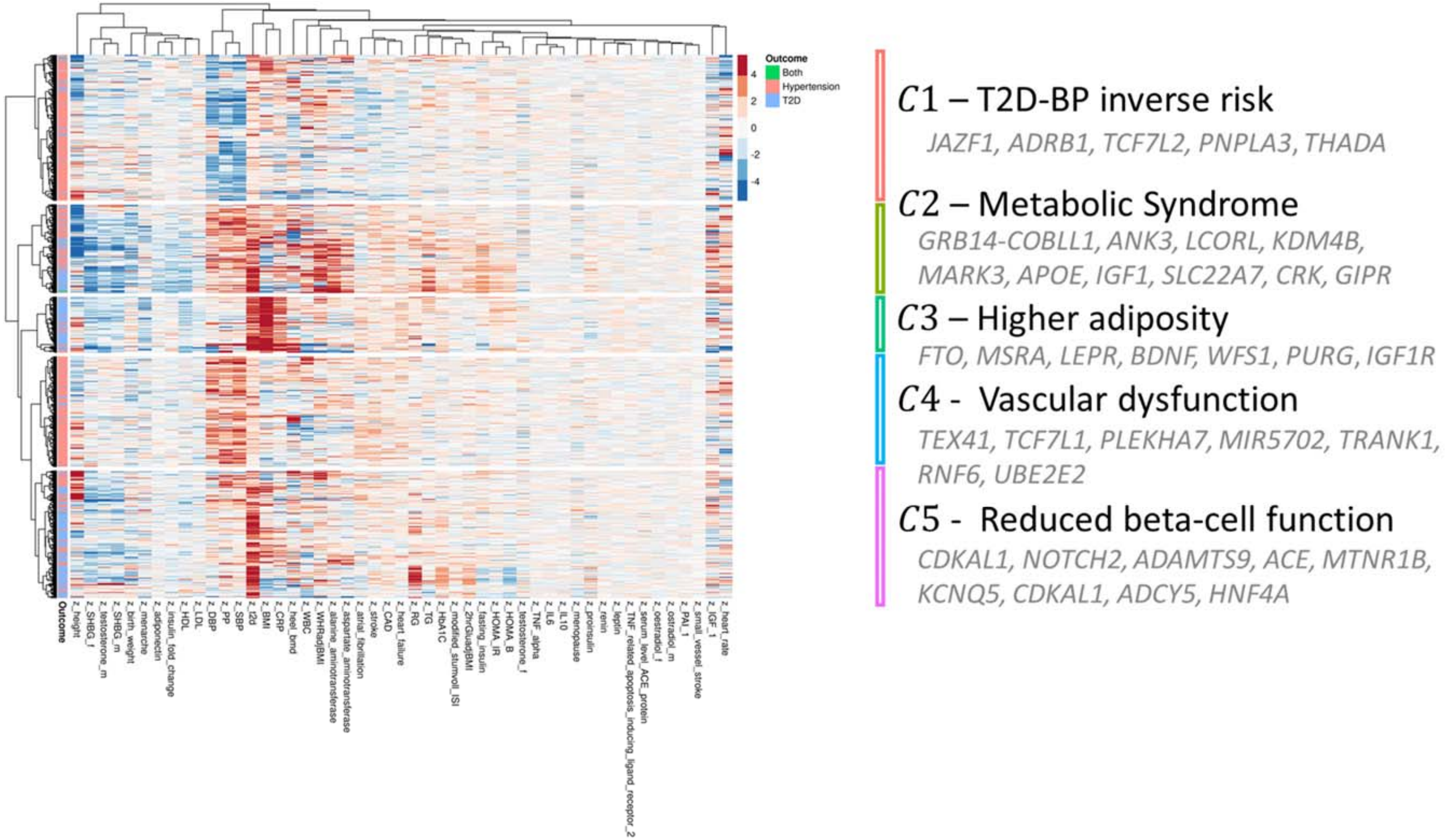


**Supplementary Figure 1:** Hierarchical clustering of the 1,304 SNVs using *pheatmap*.

Legend:

Each row is a SNV, each column is a GWAS, the effects are reported in z-scores = beta/se.
The first column represents the origin of the SNV (from T2D GWAS in blue, BP GWAS in red or both in green).
The black text to the right of the clustering is the attributed cluster names. The grey text represents associated loci within the cluster.
T2D = Type 2 Diabetes; DBP = Diastolic Blood Pressure; UKB = UK Biobank; PP = Pulse Pressure; SBP = Systolic Blood Pressure; HbA1C = Glycated hemoglobin; RG = Random glucose; WHR = waist-hip ratio; BMI = body mass index; IL = interleukin; HDL = high-density lipoprotein; PAI = Plasminogen activator inhibitor; ISI = Insulin Sensitivity Index; IGF = Insulin-like growth factor; LDL = low-density cholesterol; adjBMI = adjusted for BMI; HOMA = homeostatic model assessment; IR = insulin resistance; B = beta-cell function; WBC = white blood cell count; CRP = C-reactive protein; CAD = coronary artery disease; TG = triglycerides; SHBG = sex-hormone-binding globulin; ACE = Angiotensin-converting enzyme; TRAIL-R2 = TNF related apoptosis inducing ligand_receptor 2; ALT = alanine aminotransferase; AST = aspartane aminotransferase; BMD = bone mass density; TSH = thyroid stimulating hormone; FT4 = free thyroxine

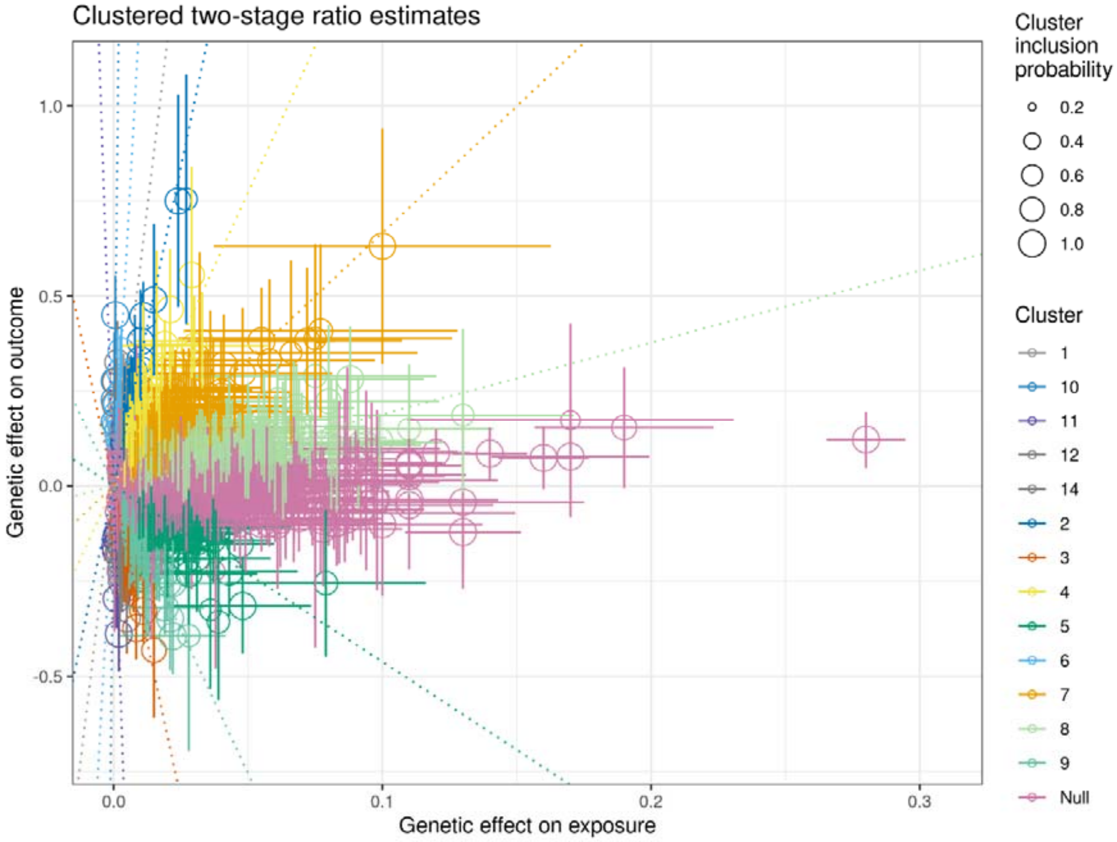


**Supplementary Figure 2:** *MRClust* results using T2D as exposure and PP as outcome using the curated 563 T2D SNVs as instrument variables (***Methods – Clusters of pathogenetic processes***).

<u>Legend:</u>

T2D = Type 2 Diabetes; PP = Pulse Pressure; SNV = single nucleotide variants; MR = Mendelian randomization

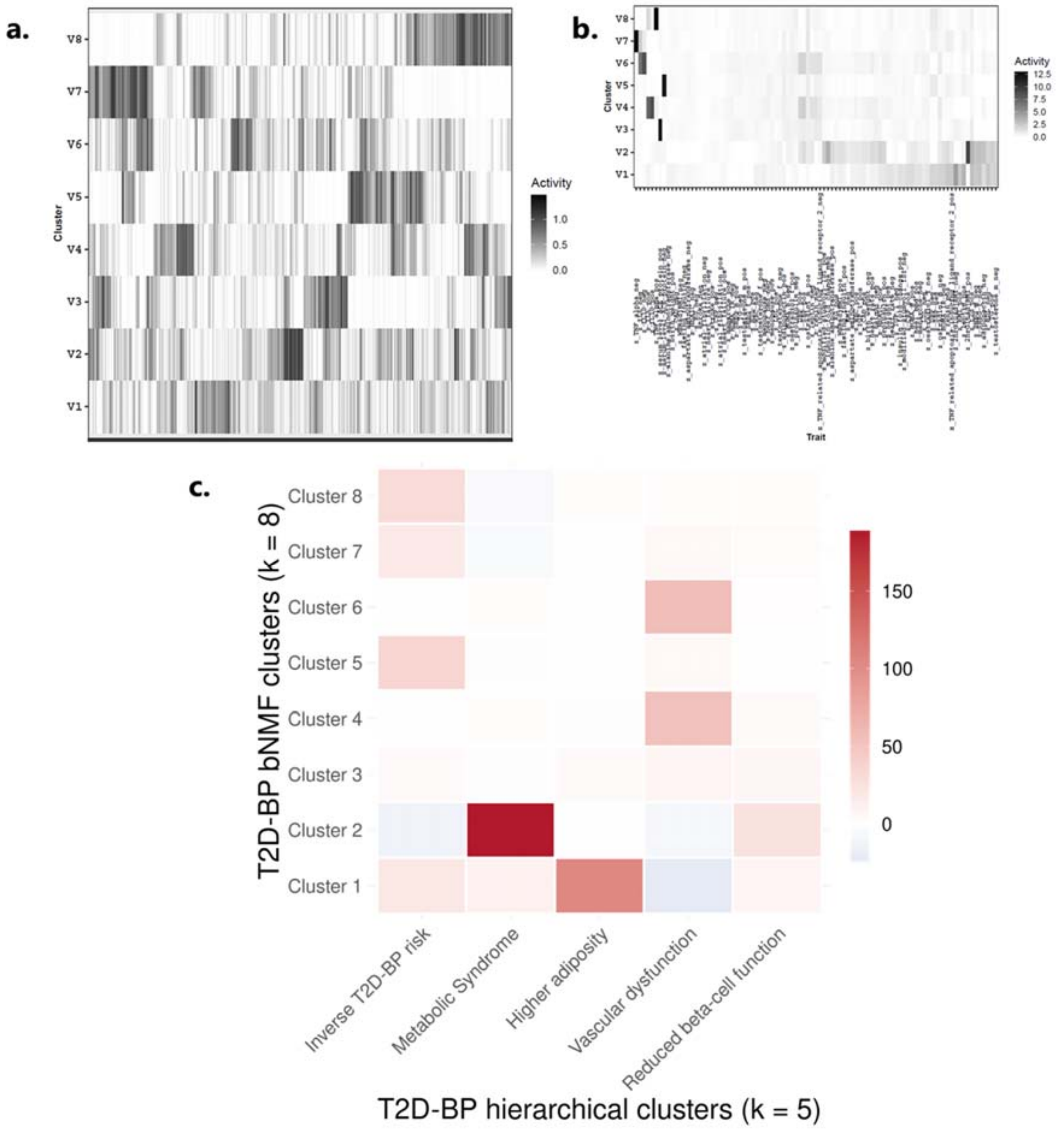


**Supplementary Figure 3:** Bayesian nonnegative matrix factorization (bNMF) clustering using the 1,304 T2D-BP SNVs, ran with default parameters and 10 iterations (***Methods – Clusters of pathogenetic processes***). Most iterations (5/10) resulted in 8 groups. **a,** Variant association to clusters. **b,** Feature (GWAS summary statistics) association to clusters. **c,** Comparison between the hierarchical clusters (columns) and the bNMF clusters (rows). Each value intensity represents the associated -log(P-value) of the linear association while the colour is the direction of the linear association beta (***Methods – Clusters of pathogenetic processes***).

<u>Legend:</u>

bNMF = Bayesian nonnegative matrix factorization; T2D = Type 2 Diabetes; DBP = Diastolic Blood Pressure; UKB = UK Biobank; PP = Pulse Pressure; SBP = Systolic Blood Pressure; HbA1C = Glycated hemoglobin; RG = Random glucose; WHR = waist-hip ratio; BMI = body mass index; IL = interleukin; HDL = high-density lipoprotein; PAI = Plasminogen activator inhibitor; ISI = Insulin Sensitivity Index; IGF = Insulin-like growth factor; LDL = low-density cholesterol; adjBMI

= adjusted for BMI; HOMA = homeostatic model assessment; IR = insulin resistance; B = beta-cell function; WBC = white blood cell count; CRP = C-reactive protein; CAD = coronary artery disease; TG = triglycerides; SHBG = sex-hormone-binding globulin; ACE = Angiotensin-converting enzyme; TRAIL-R2 = TNF related apoptosis inducing ligand_receptor 2; ALT = alanine aminotransferase; AST = aspartane aminotransferase; BMD = bone mass density; TSH = thyroid stimulating hormone; FT4 = free thyroxine

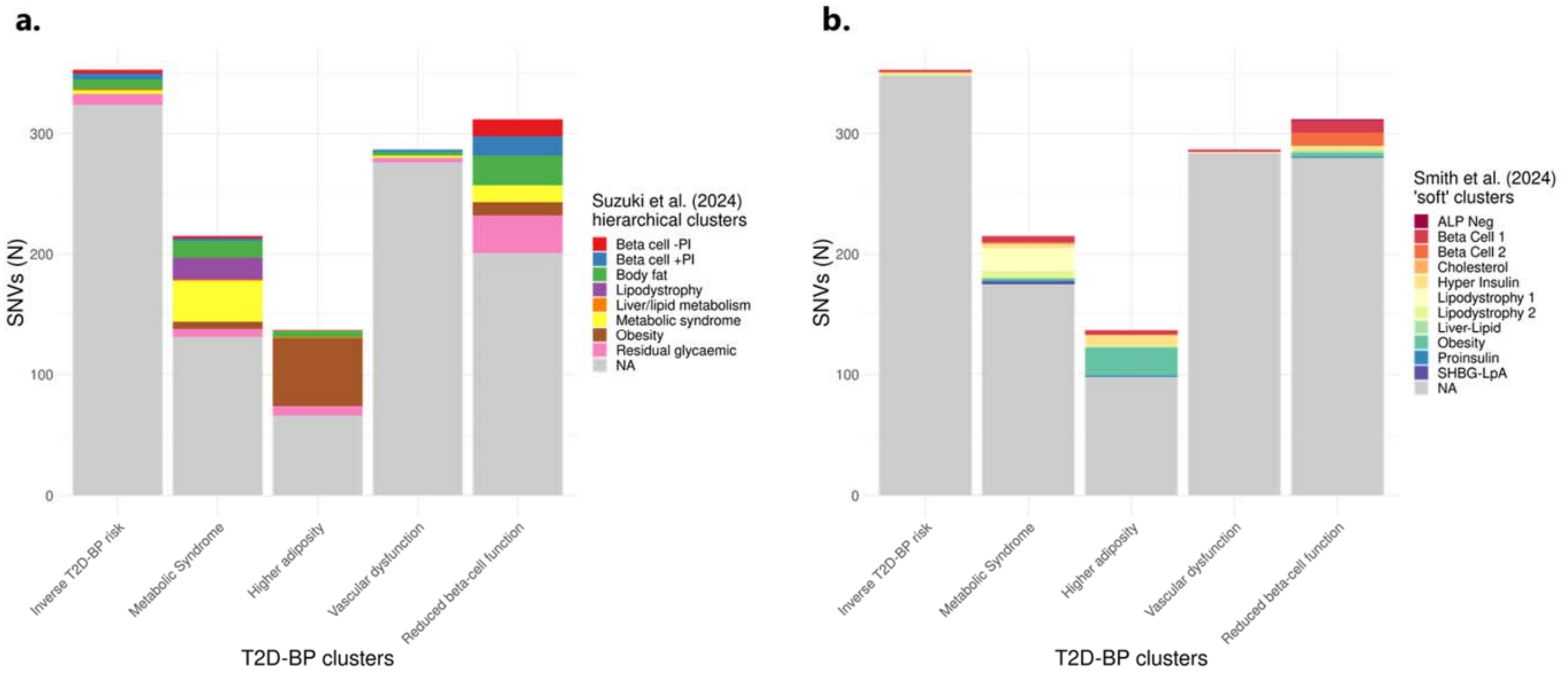


**Supplementary Figure 4: a,** SNV comparison between the T2D-BP hierarchical clusters (x-axis) and the cluster of the latest T2D hierarchical clustering (T2DGGI paper) from Suzuki et al. (2024). **b,** Comparison between the T2D-BP hierarchical clusters (x-axis) and the latest T2D 'soft' clustering from Smith et al. (2024). The comparison is done using each study genetic variants and looking for LD proxy (LD $r^2 > 0.6$) in our T2D-BP genetic variants. For 'soft' clusters, the cluster assignment is based on weight > 0.75 (***Methods – Clusters of pathogenetic processes***).

Legend:

T2D = Type 2 Diabetes; BP = Blood Pressure; PI = Proinsulin ; NA = Non attributed; ALP = alkaline phosphatase ; SHBG = sex hormone binding globulin; LpA = lipoprotein A; SNV = single nucleotide variant.

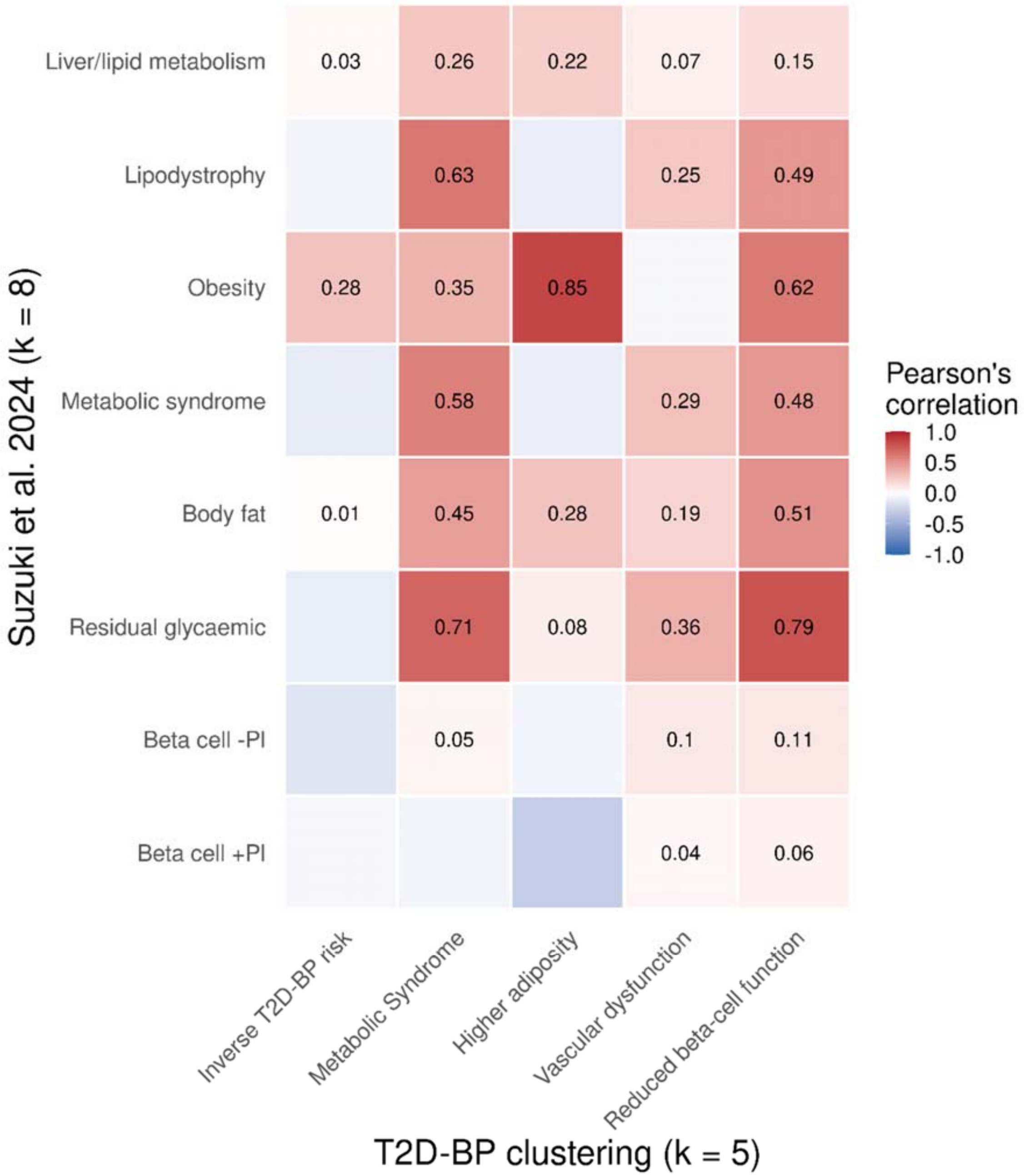


**Supplementary Figure 5:** GWAS weight comparison between the hierarchical clusters (columns) and the cluster of the T2DGGI paper (rows). Each value intensity represents the associated Pearson correlation coefficient (***Methods – Clusters of pathogenetic processes***).

Legend:

T2D = Type 2 Diabetes; BP = Blood Pressure; PI = Proinsulin

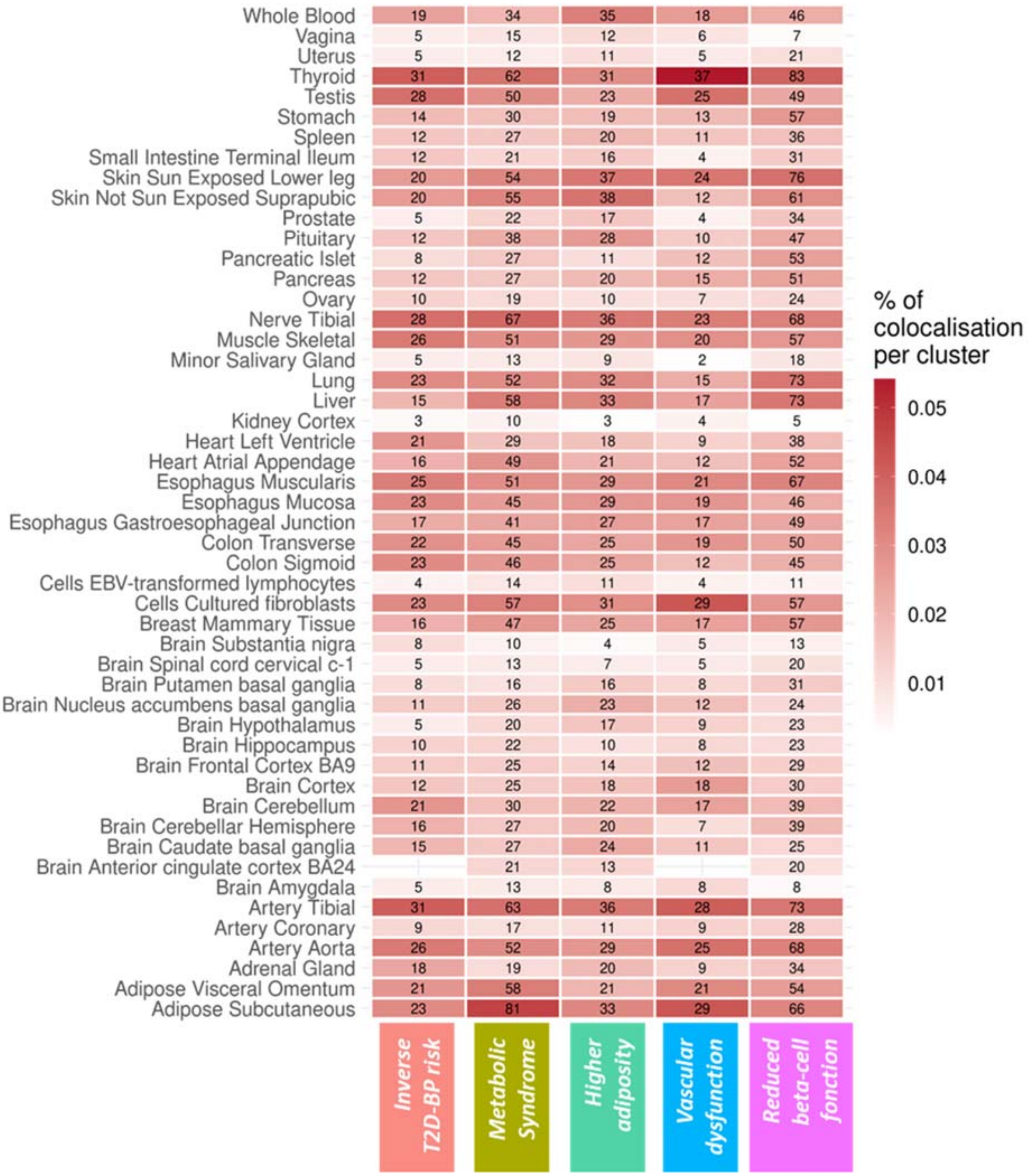


**Supplementary Figure 6:** Heat map of colocalised loci across the five clusters and 50 human adult tissues. Each column corresponds to a cluster, while each row represents a tissue. The numerical value in each tile indicates the number of colocalised loci for a specific cluster and tissue. Colour intensity corresponds to the percentage of colocalisation per cluster.

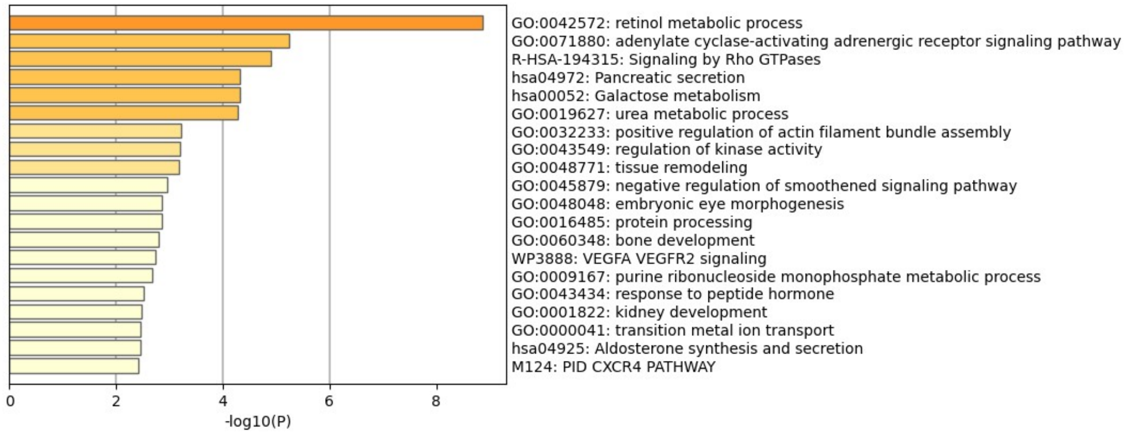


**Supplementary Figure 7:** Pathway analysis of the 202 colocalised genes within the *Inverse T2D-BP risk* cluster using *metascape*. The following genes were associated with the pathway mentioned above.

| Category | GO | Description | Colocalised genes associated |
|---|---|---|---|
| GO Biological Processes | GO:0042572 | retinol metabolic process | AKR1B1\|CYP1A1\|CYP2C8\|CYP2C18\|AKR1B10\|RDH14\|PLB1\|AKR1B15 |
| GO Biological Processes | GO:0071880 | adenylate cyclase-activating adrenergic receptor signaling pathway | ADCY9\|ADRB1\|PLN\|AKAP13 |
| Reactome Gene Sets | R-HSA-194315 | Signaling by Rho GTPases | RHOC\|ARHGAP1\|CDC20\|CSK\|PTK2\|FERMT2\|AKAP13\|SPEN\|SWAP70\|MCF2L\|NUP160\|EVL\|PLEKHG1\|EFHD2\|OBSCN\|CENPS |
| KEGG Pathway | hsa04972 | Pancreatic secretion | ADCY9\|ATP2B1\|PRSS3\|SLC4A2\|CELA2B\|CELA2A |
| KEGG Pathway | hsa00052 | Galactose metabolism | AKR1B1\|AKR1B10\|G6PC3\|AKR1B15 |
| GO Biological Processes | GO:0019627 | urea metabolic process | CYP2C9\|AGMAT\|NAGS |

| | | |
|---|---|---|
| T2D | Self-reported having T2D, verbal interview<br>OR<br>Taking medication related to T2D<br>OR<br>Diabetes diagnosed by doctor and age diabetes diagnosed > 35 and having not started insulin within the one-year diagnosis of diabetes<br>OR<br>HbA1c > 6.4%<br>OR<br>Random glucose > 11.1 mmol/l<br>OR<br>T2D is reported without T1D and gestational diabetes in ICD10 disease code | 40,619 |
| DBP | Taking the mean value of automated measurements and manual measurements<br>AND<br>Adjusting by 10 mmHg if taking blood pressure lowering medication | N/A |
| PP | SBP-DBP | N/A |
| SBP | Taking the mean value of automated measurements and manual measurements<br>AND<br>Adjusting by 15 mmHg if taking blood pressure lowering medication | N/A |
| Hypertension | SBP ≥ 150 mmHg<br>OR<br>DBP ≥ 90 mmHg<br>OR<br>Taking blood pressure lowering medication | 230,737 |

**Supplementary Figure 8:** Diagram of criteria used in the UKB to identify T2D and hypertension cases, and adjust BP levels.

Legend:

T2D = Type 2 Diabetes; BP = Blood Pressure; DBP = Diastolic Blood Pressure; UKB = UK Biobank; PP = Pulse Pressure; SBP = Systolic Blood Pressure;
